# Impacts of dynamic aerosol and pathogen concentrations on risks of *Legionella pneumophila* for Public Showers in Switzerland Based on a Quantitative Microbial Risk Assessment Framework

**DOI:** 10.1101/2025.11.20.25340595

**Authors:** Lizhan Tang, Emile Sylvestre, Kerry A. Hamilton, Frederik Hammes, Timothy R. Julian

## Abstract

*L. pneumophila* is a waterborne respiratory pathogen that causes Pontiac Fever and Legionnaires’ disease, two clinically significant diseases with increasing incidence in Europe. In this study, we develop a Quantitative Microbial Risk Assessment (QMRA) framework on the risks of infection from showering in *L. pneumophila-*contaminated water supplies to inform health-based concentration targets and water quality monitoring programs. The developed QMRA model extends on previous work investigating the relationships between concentrations of *L. pneumophila* in water sources and infection, illness, and disease burden by incorporating dynamic pathogen concentrations in water and aerosol concentrations, extending the prior reliance on assumptions of constant, average concentrations over the exposure duration. When applying this approach to data collected from within a building in Switzerland at risk for legionellosis cases, we show that initial high concentrations of *L. pneumophila* in water and aerosols from hot showers contribute to risks above a commonly used benchmark for the acceptable infection risk (10^-4^ infections per person per year) within the first 1-2 minutes of showers. Extending the model to estimate critical concentrations of *L. pneumophila* suggests concentrations at or above 2.5 × 10^3^ CFU/L to 1.6 × 10^6^ CFU/L for first draw samples and 2.5 × 10^1^ CFU/L to 1 × 10^3^ CFU/L for samples obtained after flushing would increase infection risks above the benchmark, dependent on site-specific conditions including water temperature and shower head type. These critical values align with, but are less stringent than, values reported by previous studies for showers due to our consideration of dynamic aerosol concentrations. Sensitivity analysis suggests that controlling *L. pneumophila* concentrations in water is the most effective risk mitigation strategy. Ventilation to reduce risks is dependent on shower conditions but may be less effective. The QMRA model finds that consideration of dynamic *L. pneumophila* concentrations in water improves exposure estimates and therefore improve the risk assessment, informing the benefits of sampling strategies that assess both first draw and flush samples in routine water monitoring programs.

## 1. Introduction

*Legionella* spp., the causative agent of Pontiac Fever and Legionnaires’ disease, are commonly present in engineered water systems and can be transmitted to humans through inhalation of aerosols from cooling towers and different water fixtures..^1-3^ Among *Legionella* spp., *Legionella pneumophila* is identified as the major contributor to water-associated sporadic cases as well as outbreaks.^4, 5^ An increasing trend of Legionnaires’ disease cases has been observed in recent decades in Europe, and may be exacerbated by a changing climate.^6-8^ Routine monitoring programs and management strategies for engineered water systems are recommended to minimize infection risks of *L. pneumophila*.^9, 10^

Developing effective water monitoring programs can be supported through Quantitative Microbial Risk Assessment (QMRA), which is a modeling framework that estimates risks of infection, illness, or other sequelae from exposures to environmental reservoirs. ^11-13^ QMRA generally consists of four steps, which the World Health Organization^13^ defines under a harmonized framework as: problem formulation, exposure assessment, health effects assessment and risk characterization. In problem formulation, microbial hazards are identified based on their association with specific outbreaks or sporadic cases to determine the most relevant pathogens for further risk assessments.^14^ In exposure assessment, experimental measurements are combined with mathematical modeling to describe occurrence, fate, and transport of pathogens in the environment. Combined with modeling of exposure pathways based on people’s behaviours, this step estimates the pathogen dose for modeled scenarios.^15-17^ In the health effects assessment, the relationship between dose and probability of response (e.g., risk of infection, illness, or other sequelae) is selected based on available data and informed by pathogen strain type, the host, exposure route and disease endpoint.^18-20^ Finally, in the risk characterization step, risks are annualized and converted to other measures of disease burden (e.g., disability adjusted life years [DALY]). In many cases, the estimated risk or measure of disease burden is compared with a benchmark to inform adverse health outcomes caused by the specific pathogen.^2, 21^

Existing QMRA frameworks for *L. pneumophila* are useful to inform water quality guidelines, evaluate efficiency of intervention strategies, and estimate associated waterborne disease burden. However, an underlying assumption of these QMRA models for *L. pneumophila* is that pathogen and aerosol concentrations are constant throughout the exposure event, assuming measured concentrations of *L. pneumophila* and aerosols represent steady state conditions.^2, 3, 22-25^ However, empirical studies have shown that both aerosol concentrations and pathogen concentrations are dynamic for scenarios such as faucets and showering.^26, 27^ For example, the concentration of aerosols for a shower have been shown to exponentially increase within the first few minutes before reaching steady state concentrations.^26, 28^ Similarly, variations in aerosol generation and decay rates can influence the length of time before aerosol concentrations reach steady state. In addition to the dynamism of aerosol concentrations. *L. pneumophila* water concentrations are also dynamic. Causes of variability in *L. pneumophila* concentrations include biofilm detachment and heterogeneity in concentrations throughout the piping network.^29, 30^ Several factors such as reduced hot water temperatures, longer stagnation periods, and elastomeric materials can favor biofilm and *L. pneumophila* growth at distal points in a network.^31^ This can lead to concentrations of *L. pneumophila* in the first draw samples that are substantially higher than in flush samples, especially for hot water systems.^27, 31, 32^

To better inform water monitoring and management strategies, we developed a QMRA framework for *L. pneumophila* that integrates and investigates the impact of dynamic changes of aerosol and pathogen concentrations into exposure assessment modeling. The modeling framework provides a basis for evaluating the importance of incorporating dynamic change of aerosol and pathogen concentrations on infection risks. Further, the framework provides an opportunity to evaluate the impact of current sampling strategies for regular water monitoring on risk estimates from *L. pneumophila* in showers. Finally, risk reductions from potential intervention strategies were quantified to demonstrate practical applications of the approach.

## 2. Methods

### 2.1. Exposure assessment

#### 2.1.1. Exposure scenario

We modeled exposures to showers because they generate significantly more aerosols than alternative exposure pathways, such as toilet flushing.^4^ We modeled the shower as a typical shower stall with a door and assumed a closed system with ventilation modeled at the top (ceiling) of the shower stall. For a single shower event, we assume a person will be exposed to aerosols as soon as the shower is turned on. Because we assumed the person was exposed from a single location inside the shower, this assumption may be a simplification, as people may wait outside of the shower until the water temperature stabilizes. We investigated the risk of infection for discrete time periods to evaluate the impact of this assumption on risk as further discussed. After showering, we assume a person turns off the shower but remains exposed to aerosols from the same location inside the stall for an additional 5 minutes.

#### 2.1.2. Modeling dynamic change of aerosol and *L. pneumophila* concentrations during showers

We simulated the dynamic change of aerosol concentrations for the conventional showerhead and the rain showerhead at both cold water and hot water temperature conditions to evaluate impact of water temperature on our risk assessment based on the previous work of Tang, et al. ^33^. The release of *L. pneumophila* during flushing was modeled assuming an exponential decay curve, as informed by a previous study.^30, 34^ To model the decay curve, two scenarios were assumed. First, a scenario was modeled in steady state *L. pneumophila* concentrations were assumed constant, equal to the measured value of the flush sample (i.e., worst case), and second, where influent water was assumed to be uncontaminated with *L. pneumophila* concentrations (i.e., best case). The details about the modeling of aerosol and *L. pneumophila* concentrations during flushing can be accessed in Supplemental Material Text S1 and S2.

#### 2.1.3. Temporal variability in concentrations of *L. pneumophila*

The *L. pneumophila* concentration data were obtained from six shower stalls at a school in Switzerland from November 2021 to October 2022 (Supplemental Material Table S2). The school was sampled because it was known to be an at-risk building for *L. pneumophila* contamination. The hot water for the school is produced by a water station with two heat exchangers. For the sampling, showerheads were turned on at a flow rate of approximately 5 L/min. Cold (25-30 °C) and hot water (40-60°C) samples were collected separately at each sampling point. During flushing, the 1^st^ liter samples were collected, which we refer to throughout as the first draw sample, and then the fifth liter samples were collected, which we refer to throughout as the flush sample. The enumeration of *L. pneumophila*. was conducted following ISO 11731.^35^ Isolates were also analyzed by latex agglutination test to obtain information about speciation. The *L. pneumophila* raw count data were then pooled based on the water temperature (i.e., hot water and cold water) and the sample type (i.e., first draw and flush sample).

Based on sample volumes and count data, mixed Poisson distributions were fitted to the data to characterize the temporal variability in concentrations of *L. pneumophila*, based on Sylvestre, et al. ^36^. The spatial dispersion of microorganisms in the water sample was assumed to follow a Poisson (random) distribution (equation1):

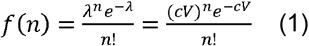

where *n* is the number of microorganisms observed in the sample; λ is the expectation of the number of microorganisms in a given sample; *c* is the unknown concentration for the sample (CFU/L); *V* is the sample volume (L).

To estimate the variability in sample concentration *c*, we used either the lognormal distribution or the gamma distribution as these distributions represent approximations of growth models under varying assumptions of stable equilibrium and environmental carrying capacity.^37^ The probability density function for a lognormal distribution is:

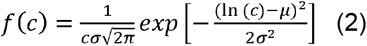

where μ is the mean of the natural logarithm of unknown concentration *c*; *σ* is the standard deviation of the natural logarithm of unknown concentration *c*.

The probability density function for a gamma distribution is:

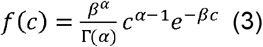

where *α* is the shape parameter; *β* is the scale parameter.

Bayesian inference with Markov Chain Monte Carol (MCMC) was applied to obtain parameter values for mixed Poisson distributions. Uninformed prior distributions were set for the parameters of the lognormal and gamma distributions to minimize the impact of priors on posterior distributions. For the gamma distribution, the prior distribution for shape was set as uniform (0,1) and for rate as uniform (1×10^-12^, 0.1). For lognormal distributions, the prior distribution for mean was uniform (0,1) and standard deviation was exponential (1). Marginal deviance information criterion (mDIC) values were used to compare the fit of the distributions to empirical data.^36^ Lower mDIC values indicate better fit and a difference of 3 in mDIC was considered significant for model selection.^38^

#### 2.1.4. Modeling the association between first draw and flush concentrations

Within the data set of paired first draw and flush samples, we observed a positive association between the first draw sample concentrations and flush sample concentrations. Since the data were recorded as counts, we modelled this relationship using a negative binomial generalized linear mixed effect regression model, which accounts for the overdispersion typically present in microbial count data. The model predicts the *L. pneumophila* count in the flush sample based on the first draw count, using a log-link function and including a random intercepts to account for between-shower variability. The model is specified as follows:

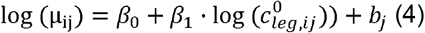

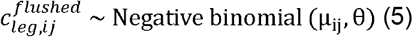

where 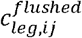 is the *L. pneumophila* count in the flush sample from shower unit *j*, observation *i*, 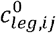 is the *L. pneumophila* count in the corresponding first draw sample, *β*_1_ is the fixed effect, *β*_*0*_ is the model intercept, *b*_*j*_ is the random effect for shower unit *j* that follows a normal distribution, *μ* is expected value of the flushed count and *θ* is the shape (dispersion) parameter for the negative binomial distribution.

We were interested in the estimated coefficient *β*_1_, which quantifies the association between *L. pneumophila* count in first draw and flush samples. A significantly positive *β*_1_ indicates that higher counts in first draw samples are associated with higher counts in corresponding flush samples, after accounting for between-shower variability and overdispersion.

#### 2.1.5. Cumulative dose estimation

The cumulative dose (CFU) for a shower can be estimated as follow.

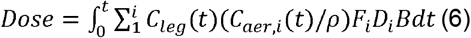

Where *C*_*leg*_*(t)* is the concentration of *L. pneumophila* at time t (CFU/L water); *C*_*aer,i*_*(t)* is the concentration of aerosols of size class i at time t (g/m^3^ air); *ρ* is the density of water (g/L water); *B* is the inhalation rate (m^3^ air/min); *F*_*i*_ is the fraction of *L. pneumophila* that partitions to aerosols of size class i relative to bulk water; *D*_*i*_ is the deposition efficiency of aerosols of size class i at alveoli.

#### 2.1.5. Dose-response relationships

The exponential dose-response model was applied for subclinical infection by *L. pneumophila*.^18^

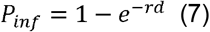

Where *P*_*inf*_ is the probability of infection for a single exposure event; *r* is the independent probability of surviving and initiating infection for each *L. pneumophila* that reaches the alveoli (/CFU); *d* is the dose for a single exposure event (CFU).

The probability of illness for different demographic groups can be estimated based on morbidity ratio proxy for children, adults and elderly.^39^

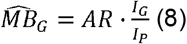

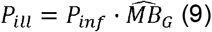

Where *AR* is the attack rate, *I*_*G*_ is the incidence rate for the demographic group, *I*_*P*_ is the incidence rate for total Swiss population, *P*_*ill*_ is the probability of illness; 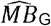 is the morbidity ratio proxy estimated from Legionnaire’s disease incidence rate and the national attack rate for specific demographic groups.

#### 2.1.6. Risk characterization

Annual risk of infection and risk of illness were calculated as follow.

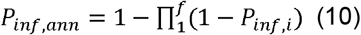

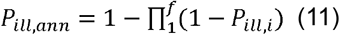

Where *P*_*inf,ann*_ is the annual risk of infection; *P*_*ill,ann*_ is the annual risk of illness; *f* is the number of showers per year, *P*_*inf,i*_ is the probability of infection for shower *i*, sampled from the distribution of *P*_*inf*_, and *P*_*ill,i*_ is the probability of infection for shower *i*, sampled from the distribution of *P*_*ill*_.

The disability adjusted life years (DALY) per case of Legionnaire’s disease was calculated based on years lost due to death (YLL) and years lost due to disability (YLD).^40^

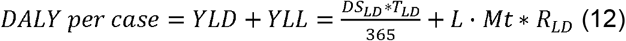

Where *DS*_*LD*_ is the disease severity for legionnaire’s disease; *T*_*LD*_ is the duration for Legionnaires’ disease (d); *R*_*LD*_ is the percentage of Legionnaire’s disease cases among total legionellosis cases (%); *L* is the years of life lost due to early death amongst Legionnaire’s disease patients (years); *Mt* is the mortality rate for Legionnaire’s disease patients (%). Assumed parameter values are available in Table S1.

The annual DALY burden was then calculated as follows.

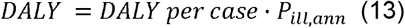

#### 2.1.7. Model implementation and sensitivity analysis

All analyses were performed in R (v4.4.1) and RStudio (v2024.04.2). The differential equations for dynamic change of aerosol concentrations, *L. pneumophila* concentrations and cumulative dose were solved using the ode.1D function in deSolve.^41^

Bayesian inference with Markov Chain Monte Carol (MCMC) was applied to obtain parameter values for mixed Poisson distributions with rjags (v4-10).^42^ For each parameter, three Markov chains were run for 10^5^ iterations after a burn-in phase of 10^3^ iterations. The Brooks–Gelman– Rubin scale reduction factor was used to evaluate the convergence of the chains. The effective sample size (the ratio of the sample size to the amount of autocorrelation in the Markov chains) was evaluated to ensure that the full posterior distribution was explored. A value of effective sample size higher than 10000 was considered sufficient for full exploration. Brooks–Gelman– Rubin scale reduction factor and effective sample size were calculated using diagMCMC function.^43^

A global sensitivity analysis was carried out to identify the contribution of parameter variability and uncertainty to the model output, which was specified as the daily risk of infection. For the sensitivity analysis, similar results were obtained using 10^3^ and 10^4^ Monte Carlo iterations with seed value set at 100. Therefore, 10^3^ iterations were used to reduce model running time. The Spearman rank correlation coefficient was used to identify the relative contributions of parameter estimates on the variation in the outcome.

In addition, a local sensitivity analysis was conducted. To determine the impact of a single parameter on model outcomes, one parameter was varied within a range while all other parameters were kept constant at their median value. To conduct the alternative scenario modeling, we assumed an initial *L. pneumophila* concentration of 10^5^ CFU/L and steady state *L. pneumophila* concentration of 10^4^ CFU/L for cold water showers. For hot water showers, the initial *L. pneumophila* concentration was 10^3^ CFU/L and steady state *L. pneumophila* concentration was 10^2^ CFU/L. Ventilation rate was assumed to be 0.7 m^3^/min. All other model parameter values were the same as the values we applied for the QMRA (Table S1). The sensitivity of the parameter can be expressed as follows:

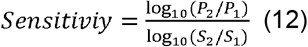

Where *P*_*1*_ is the value of infection risk at baseline condition; *P*_*2*_ is the value of infection risk at the new condition; *S*_*1*_ is the value of studied parameter at baseline condition; *S*_*2*_ is the value of studied parameter at the new condition.

## 3. Results

### 3.1. Distribution of *L. pneumophila* concentrations in showers

For first draw sample concentrations, Poisson–gamma distributions better described the variation in the samples than Poisson-lognormal distributions based on the estimated mDIC values and visualization of the behavior of the upper tail distributions (Figure 1). We note Figure 1 visualizes the probability of exceeding a given concentration based on Poisson-gamma or Poisson-lognormal distributions. The model predicted *L. pneumophila* concentrations were generally consistent with *L. pneumophila* concentrations estimated from the count and volume data (Table 1).

**Table 1.** Summary of measured and model predicted concentrations of *L. pneumophila* (CFU/L) in shower water for pooled results from first draw and flush samples from both the cold and hot water lines from six showers sampled over 1 year from a building in Switzerland with known *L. pneumophila* contamination.

| Sample type | Cold Water |  |  |  | Hot Water |  |  |  |
| --- | --- | --- | --- | --- | --- | --- | --- | --- |
|  | First Draw |  | Flush Samples |  | First Draw |  | Flush Samples |  |
|  | Measured | Model predicted | Measured | Model predicted | Measured | Model predicted | Measured | Model predicted |
| Arithmetic mean | 11780 | 11888 | 3940 | 3204 | 3191 | 3095 | 1529 | 1286 |
| Median | 6000 | 6603 | 970 | 2639 | 680 | 1048 | 200 | 858 |
| Standard deviation | 14985 | 14762 | 6349 | 2524 | 5921 | 5180 | 3117 | 1345 |
| No. of samples | 38 |  | 38 |  | 37 |  | 37 |  |

**Figure 1.**
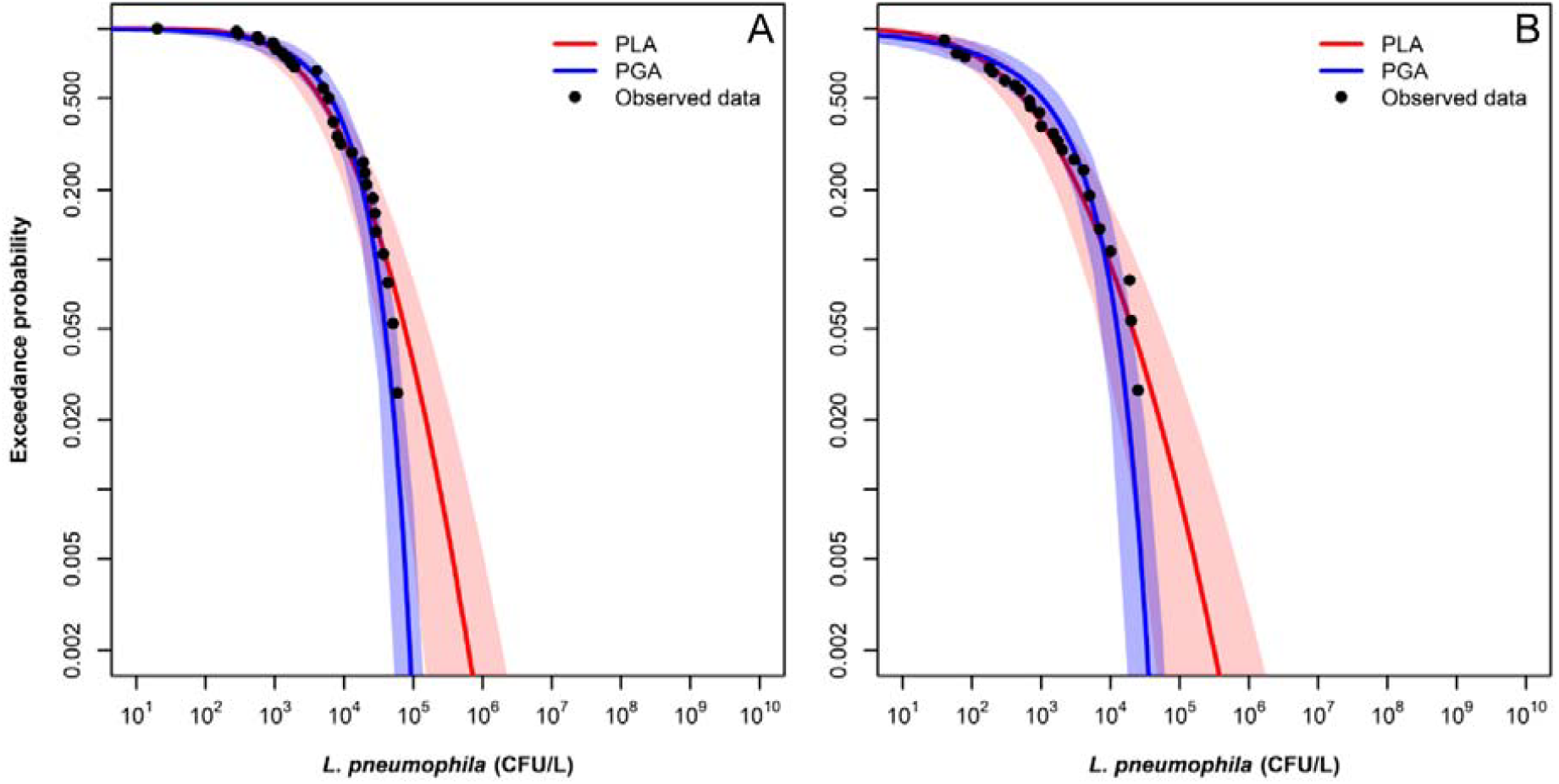
Complementary cumulative distribution function (CCDF) of distributions of *L. pneumophila* concentrations in first draw samples (CFU/L) for (A) cold water and (B) hot water. The shaded areas represent 95% confidence intervals. PGA refers to Poisson-gamma distribution (blue line and shading) and PLN refers to Poisson-lognormal distribution (red line and shading). The mDIC for cold water PGA is 730 and for PLN is 2493, and the mDIC for hot water PGA is 586 and for PLN is 2494,suggesting better fit with PGA for both cold and hot water.

Flush sample concentrations were predicted based on the first draw sampling using a fitted negative binomial regression model. Initially, the variance and standard deviation for the random effect (shower unit ID) were 1.6 × 10^-4^ and 1.2 × 10^-2^ which suggested that variability among showers was smaller than variability within showers (Figure S1 and S2). Therefore, the random effect was removed from the final regression model. The correlation coefficient describing the linear relationship between first draw sample count and flush sample count was 0.61 with a standard error of 0.07 (p = 2×10^-16^, Figure S3). The concentrations in cold water were generally higher than in hot water samples (Figure S4).

### 3.2. Risk of infection from showering in building

Both water temperature and showerhead type influenced infection risks from showering, with the scenario leading to the highest risk being the hot water shower with a conventional showerhead. Higher risks were observed at hot water temperature than cold water temperature even though the bacteria concentrations in cold water samples were higher (Figure 2). This is due to greater aerosol generation rates at hot water temperatures than cold water temperatures (Table S1). For the scenario presented in Figure 2, it is assumed that the *L. pneumophila* concentration in water after the fifth liter is constantly equal to the flush concentration until the end of the shower. Based on a benchmark of 10^-4^ infections per year, a commonly used benchmark for the acceptable burden^44^, the median annual risk of infection for a shower with cold water and a rain showerhead remained below the benchmark, while median annual risk for conventional showerhead exceeded the benchmark approximately halfway through the shower (Figure 2). For hot water showers, due to the observed rapid increase in, and subsequent high concentration of, aerosols in the first few minutes^33^, the cumulative annual risks of infection exceeded the benchmark within the first minute (Figure 2). In the scenario where the *L. pneumophila* concentration in water after the fifth liter is assumed to be constantly equal to zero until the end of the shower, the cumulative infection risks at hot showers still exceeded the benchmark within the first minute (Figure 3). This further indicates the contributions of initial high concentrations of aerosols and *L. pneumophila* to the risks at hot water temperatures. To annualize risk, we modeled *L. pneumophila* concentration profiles for each day independently. Assuming the same *L. pneumophila* concentration profiles every day when annualizing risks can lead to similar central tendency (i.e., similar mean and median results) but increases the range of scenarios (i.e., associated uncertainty and variability) (Figure S5).

**Figure 2.**
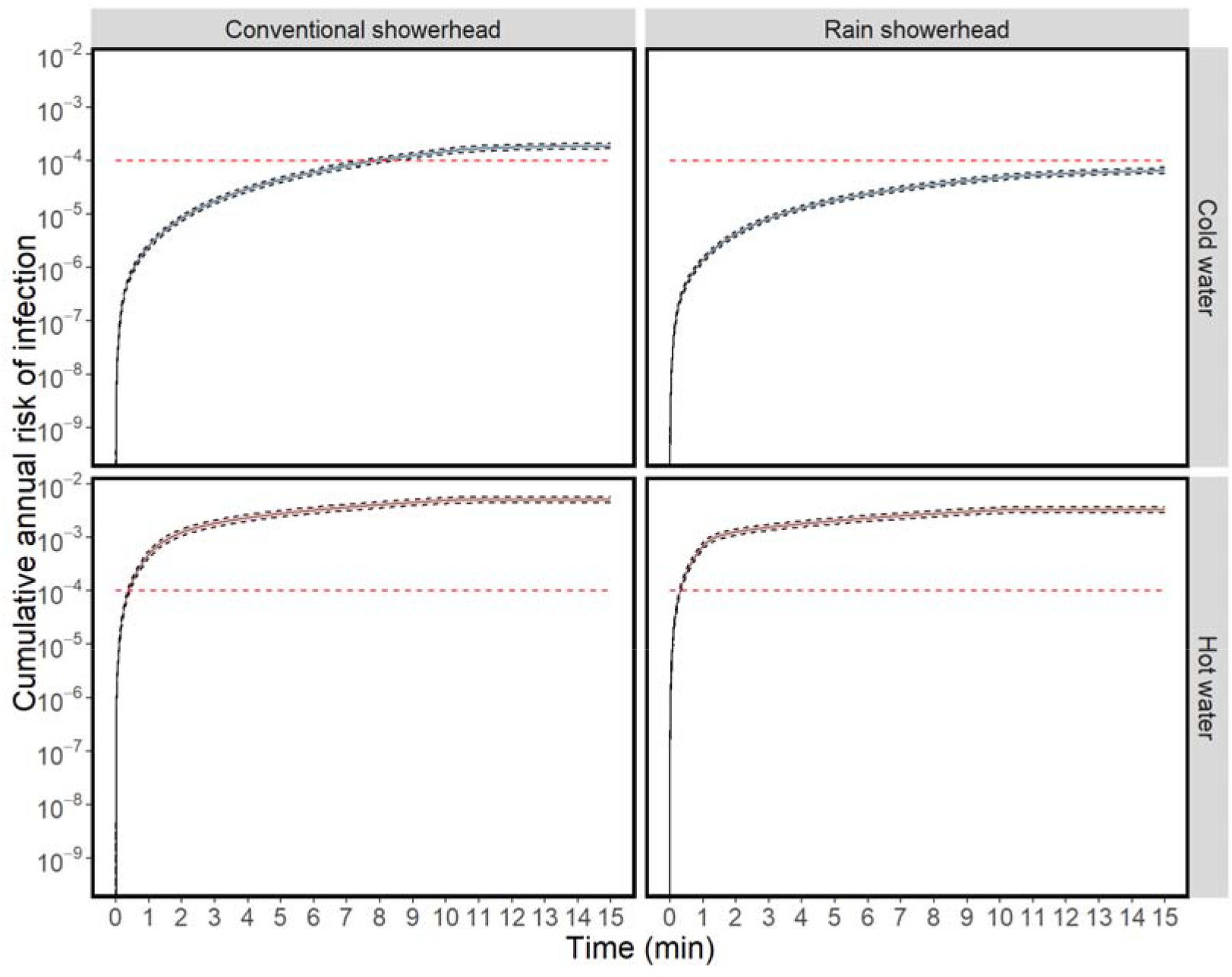
Cumulative annual risks of infection during a typical 15-minute shower based on empirical measurements from a public shower in Switzerland assuming that the initial *L. pneumophila* concentration in water is equal to the first draw sample concentration and then the concentration decays to a baseline concentration represented by the flush concentration. Solid black lines represent median risks. Black dashed lines represent 2.5% percentile and 97.5% percentile values of predicted risks. Shaded areas represent the 95% uncertainty intervals. Red dashed horizontal lines reflect the risk benchmark of 10^-4^ infections per year.

**Figure 3.**
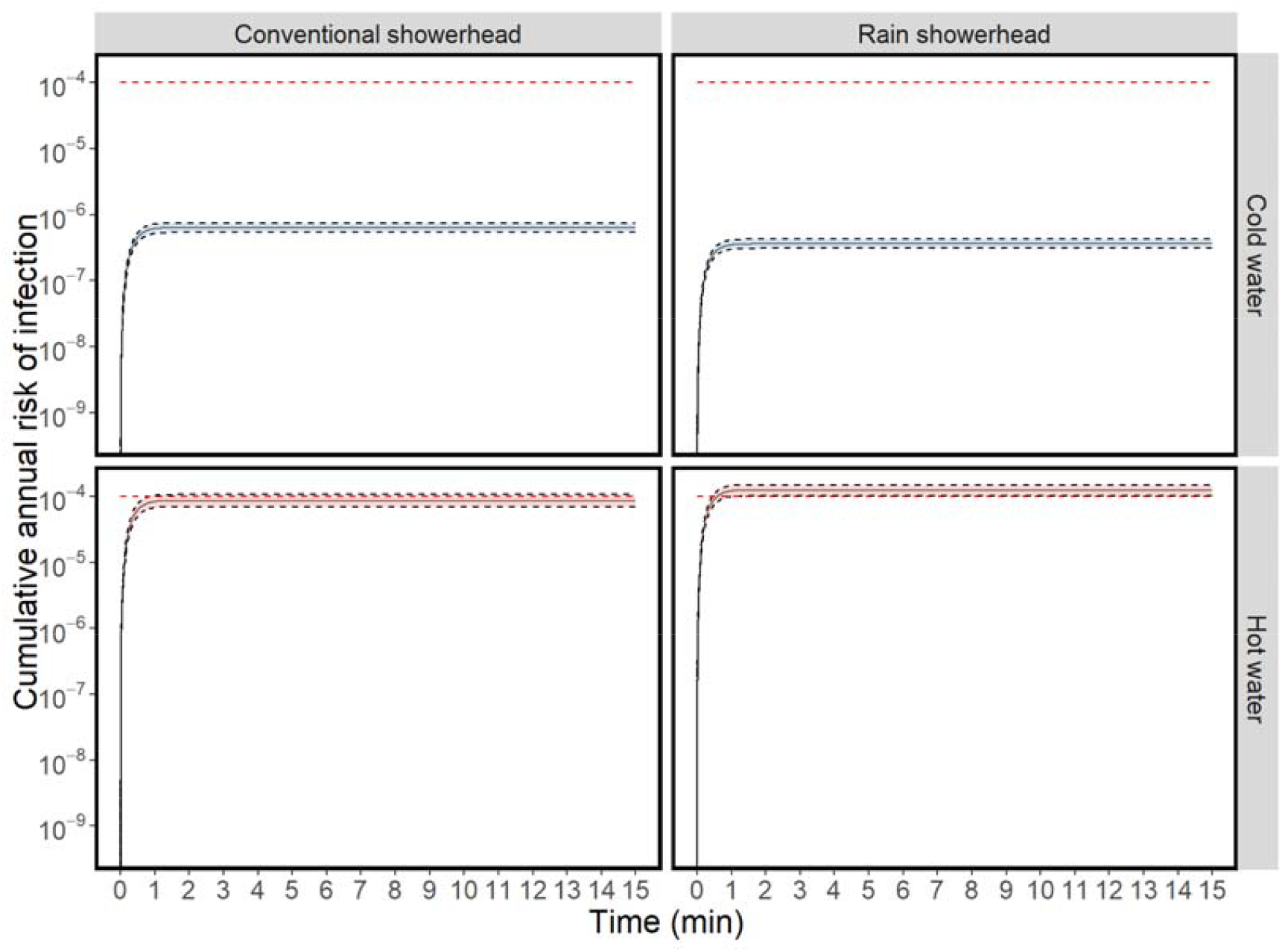
Cumulative annual risks of infection during a typical 15-minute shower based on empirical measurements from a public shower in Switzerland assuming that the *L. pneumophila* concentration in water after the first liter decaysto 0. . Solid black lines represent median risks. Black dashed lines represent 2.5% percentile and 97.5% percentile values of predicted risks. Shaded areas represent the 95% uncertainty intervals. Red dashed horizontal lines reflect the risk benchmark of 10^-4^ infections per year.

To better understand dynamic change of risks during showers, risks of infection for each 1-minute period for adults were estimated (Figure S6 and S7. The median risk of infection for each 1-minute period in cold water showers increased slightly while the shower was on, reaching a peak median risk of 2.4 × 10^-5^ for a conventional showerhead and 6.4 × 10^-6^ for a rain showerhead, well below the risk benchmark of 10^-4^ (Figure S6). Comparatively, median risks at hot water temperature were substantially higher, even in the first minute, reaching a peak risk of 7.6 × 10^-4^ for conventional showerhead and 7.0 × 10^-4^ for rain showerhead, always above the risk benchmark until showers were turned off (Figure S7). Notably, for hot showers, the risks for the first and second minutes were higher than other periods due to the empirically observed peak concentrations of aerosols.^33^ After turning off showers, a faster decrease of risks was observed at the hot water temperature than the cold water temperature due to faster decay rates of aerosol concentrations (Table S1).

Morbidity ratios were used to adjust infection risks for different demographic groups of children, adults, and the elderly (Figure 4). For the worst case scenario, the median risks for adults taking hot showers were 6.3 × 10^-5^ for a conventional showerhead and 4.1 × 10^-5^ for a rain showerhead. The median risks for the elderly taking hot showers were 1.9 × 10^-4^ for a conventional showerhead and 1.3 × 10^-4^ for a rain showerhead. Risks from cold showers were much lower, and for most demographic groups and showerhead types were below 10^-5^. Children were at the least risk compared to other groups, with median risks ranging from 2.1 × 10^-8^ to 1.7 × 10^-6^.

**Figure 4.**
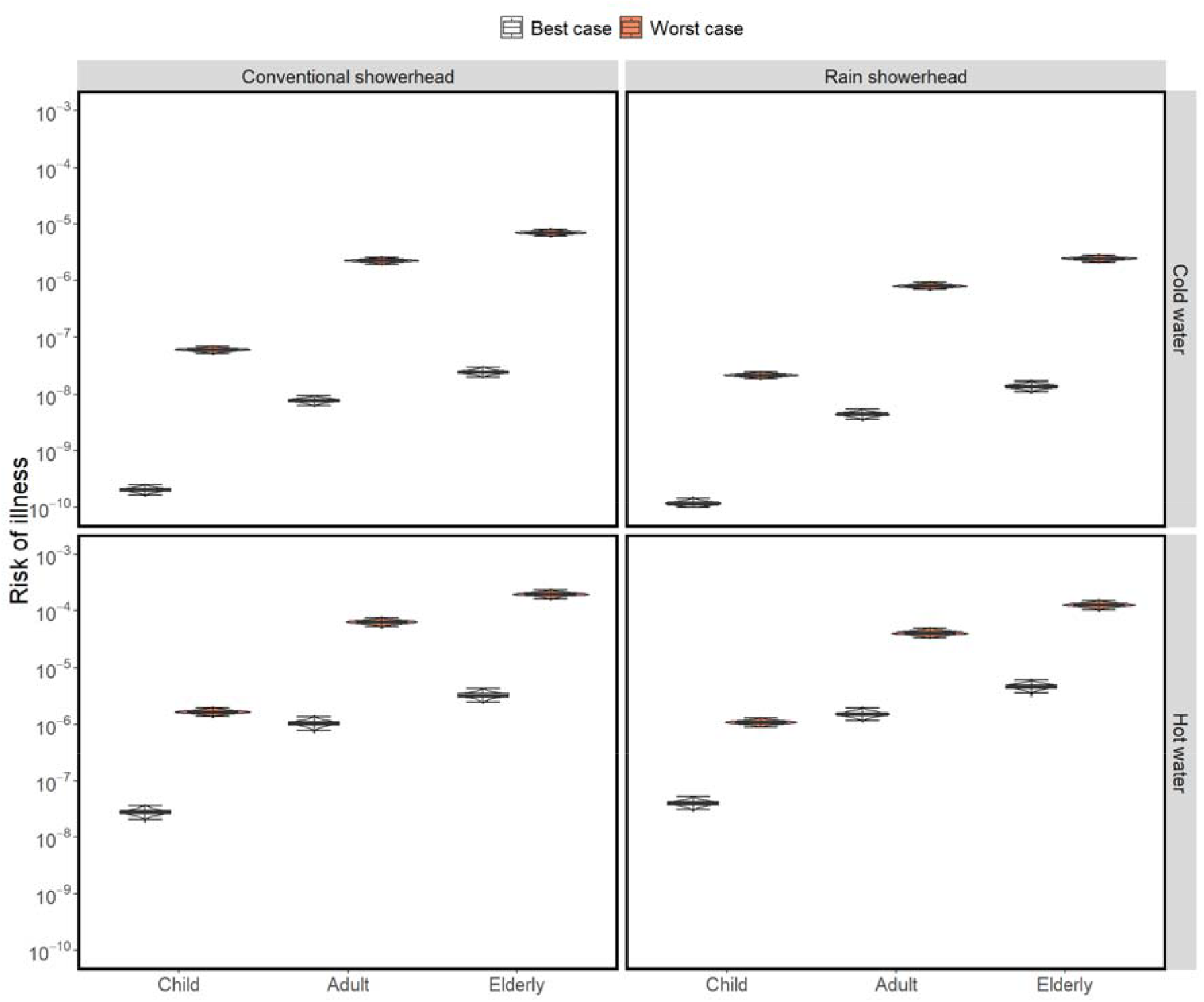
Annual risks of illness for children, adults, and the elderly for conventional and rain showers at cold and hot water temperatures based on *L. pneumophila* concentrations measured at a school in Switzerland. The worst case scenario assumes that the *L. pneumophila* concentration in water decays from the concentration measured in the first draw sample to to the flush concentration until the end of the shower and the best case scenario assumes that the *L. pneumophila* concentration in water after the first liter decays to the 0. Boxplots represent the median and interquartile range with whiskers representing the minimum to maximum range.

For the same scenario, extending the risks of infection to estimated DALYs for Legionnaire’s disease at different shower conditions, the median DALYs estimated for adults and the elderly taking hot showers exceeded the standard DALY benchmark of 10^-6^ DALYs per person per year, ranging from 4.2 × 10^-6^ for a cold water rain showerhead to 2.0 × 10^-5^ for a hot water conventional showerhead (Figure 5). The risks of illness and DALYs for the best case scenarios were 5 orders of magnitude lower at cold water temperatures and 3–4 orders of magnitude lower at hot water temperatures than the worst case scenarios.

**Figure 5.**
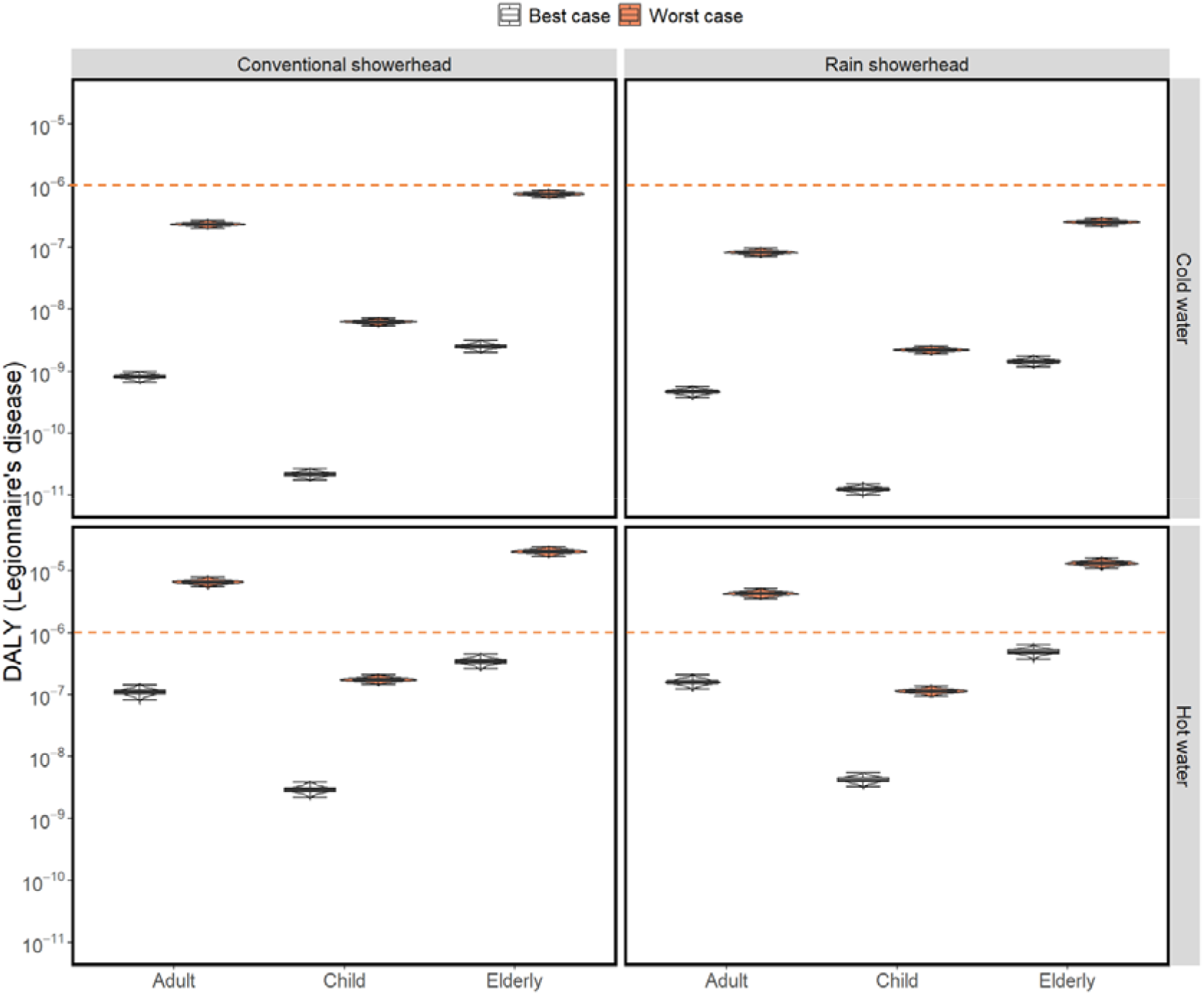
Estimates of the disability adjusted life years (DALYs) for selected shower conditions based on measurements of *L. pneumophila* in public showers at a school in Switzerland. The worst case scenario assumes that the *L. pneumophila* concentration in water decays from the concentration measured in the firs draw sample to the flush concentration until the end of the shower and the best case scenario assumes that the *L. pneumophila* concentration in water after the first liter decays to the 0. Boxplots represent the median and interquartile range with whiskers representing the minimum to maximum range. The red horizontal dashed line represents the risk benchmark of 10^-6^ DALY per year.

### 3.3. Critical concentrations of *L. pneumophila* leading to exceedances of risk benchmarks

Both first draw concentrations and flush concentrations influence infection risk estimates, as shown for conventional showerheads (Figure 6). The simulations were conducted based on scenarios using conventional showerheads as they produce a greater number of aerosols than rain showerheads (Table S1). For first draw samples, concentrations larger than 1.6 × 10^6^ CFU/L in cold water or 2.5 × 10^3^ CFU/L in hot water can lead to infection risks above the benchmark (>10^-4^ per person per year). Similarly, for flush samples, concentrations larger than 10^3^ CFU/L for cold water or 2.5 × 10^1^ CFU/L for hot water can lead to infection risks above the risk benchmark. The critical concentrations for flush samples were higher than first draw samples because flush sample concentrations contribute to *Legionella pneumophila* in aerosols for most of the shower duration. Risks are higher for hot water than cold water due to the increase in aerosol generation during showers. Based on the benchmark values, most of the observed hot water samples collected (85%) indicated risks above the benchmark, while only half of the cold water samples indicated significant risks (Figure 6). Median critical concentrations estimated assuming constant *L. pneumophila* concentrations during showers were 1905 CFU/L with a 95% confidence interval of [661, 6026] CFU/L for cold showers and 29 CFU/L with a 95% confidence interval of [10, 91] CFU/L for hot showers (Figure S8).

**Figure 6.**
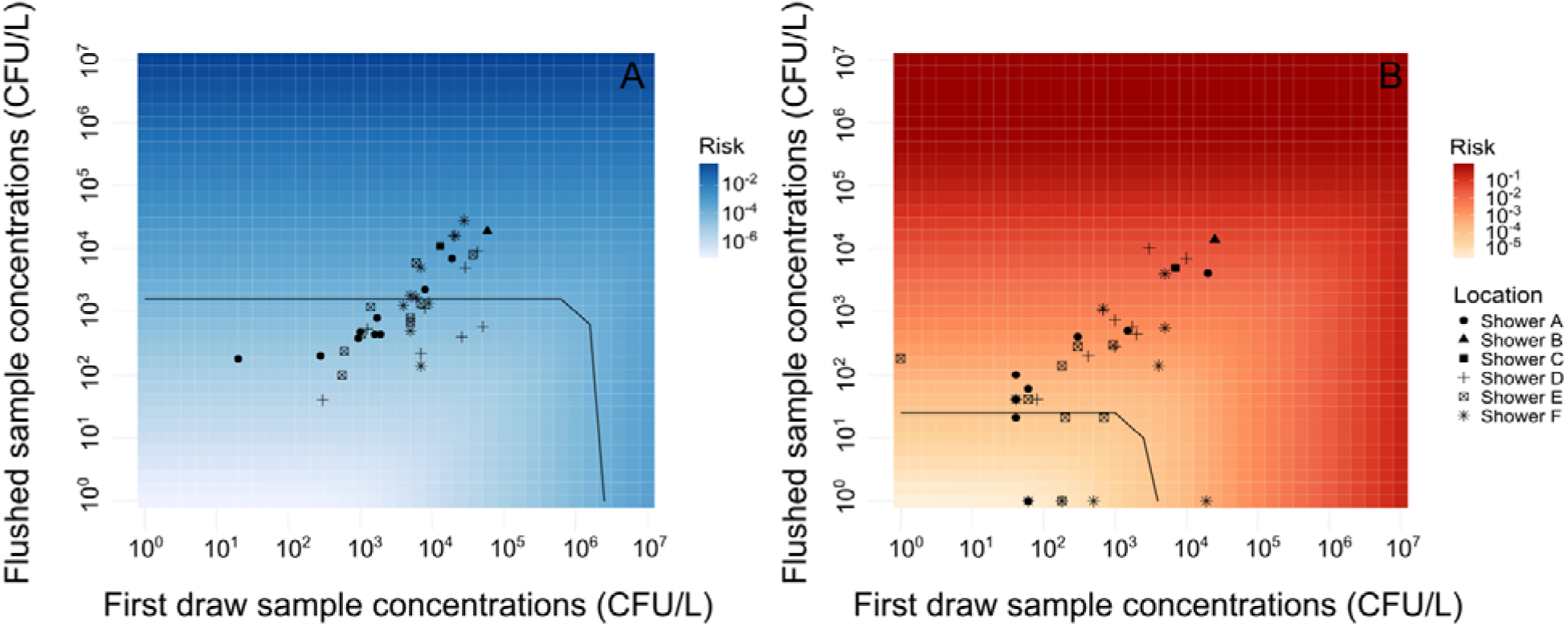
Heatmaps of the infection risk associated with showering for combinations of first draw concentrations (CFU/L) and flush concentrations (CFU/L) for (A) cold water and (B) hot water showers. The intensity of color indicates median risks of simulations. Black lines represent a risk benchmark of 10^-4^ infections per person per year. Colored data points represent empirically measured results from shower samples collected at different locations within the school in Switzerland.

### 3.4. Influential model parameters

The Spearman rank correlation coefficients indicated that the concentration of *L. pneumophila* (ρ=0.93-0.97) and the dose-response parameter (ρ=0.28-0.29) contributed substantially to the variability and uncertainty of predicted risks (Figure S9 and Figure S10). This is consistent with previous QMRA studies that also identified these two parameters as the most influential paramters.^2, 39, 45^ Comparatively, the aerosol generation rates, decay rates, as well as ventilation rates were less influential parameters (ρ=0-0.1). Previous studies applying volumetric estimation methods identified concentration of aerosols as one of the most important inputs that can influence risk outcomes.^3, 46, 47^ The difference in the results is driven by how we applied parameters to describe fate and transport of aerosols beyond applying aerosol concentrations as direct inputs. The uncertainty and variability of aerosol concentration was contributed by all aerosol transport related parameters such as aerosol generation rate, ventilation rate and other environmental conditions. Therefore, it is reasonable that aerosol concentration contributed to larger uncertainty to model outputs than any individual aerosol related parameter.

In addition to investigating the contribution of variability and uncertainty of model parameters, the impact of parameters on predicted risks can be evaluated based on alternative scenario modeling (Figure 7). The estimated baseline median infection risk estimate is 2.4×10^-4^ infections pppy for the cold shower and 2.3×10^-4^ infections pppy for the hot shower (Figure 7). Both cold and hot showers showed that reducing steady state concentrations of *L. pneumophila* was an effective way for risk mitigation, with 70% decrease of the baseline concentrations could reduce risks to a safe level (<10^-4^). Other parameters such as aerosol generation rates and shower frequency could also potentially be influential but substantial reductions nevertheless fail to reduce the risks to the benchmark level. We note that reducing first draw concentrations of *L. pneumophila* minimally impacts risks. This is influenced by our baseline assumptions, in which we applied high steady state *L. pneumophila* concentrations that already lead to risks above the benchmark. Alternative baseline assumptions may lead to differing results. We also observed that the ventilation rate had larger impact on risks for cold showers than hot showers (Figure 7). Ventilation process dominates aerosol removal at cold water temperature while other aerosol removal processes are responsible for removal at hot water temperatures. Increasing shower stall volume, which dilutes aerosol concentrations due to assumptions of well-mixed air, was more effective for hot showers than increasing ventilation (Figure 7B).

**Figure 7.**
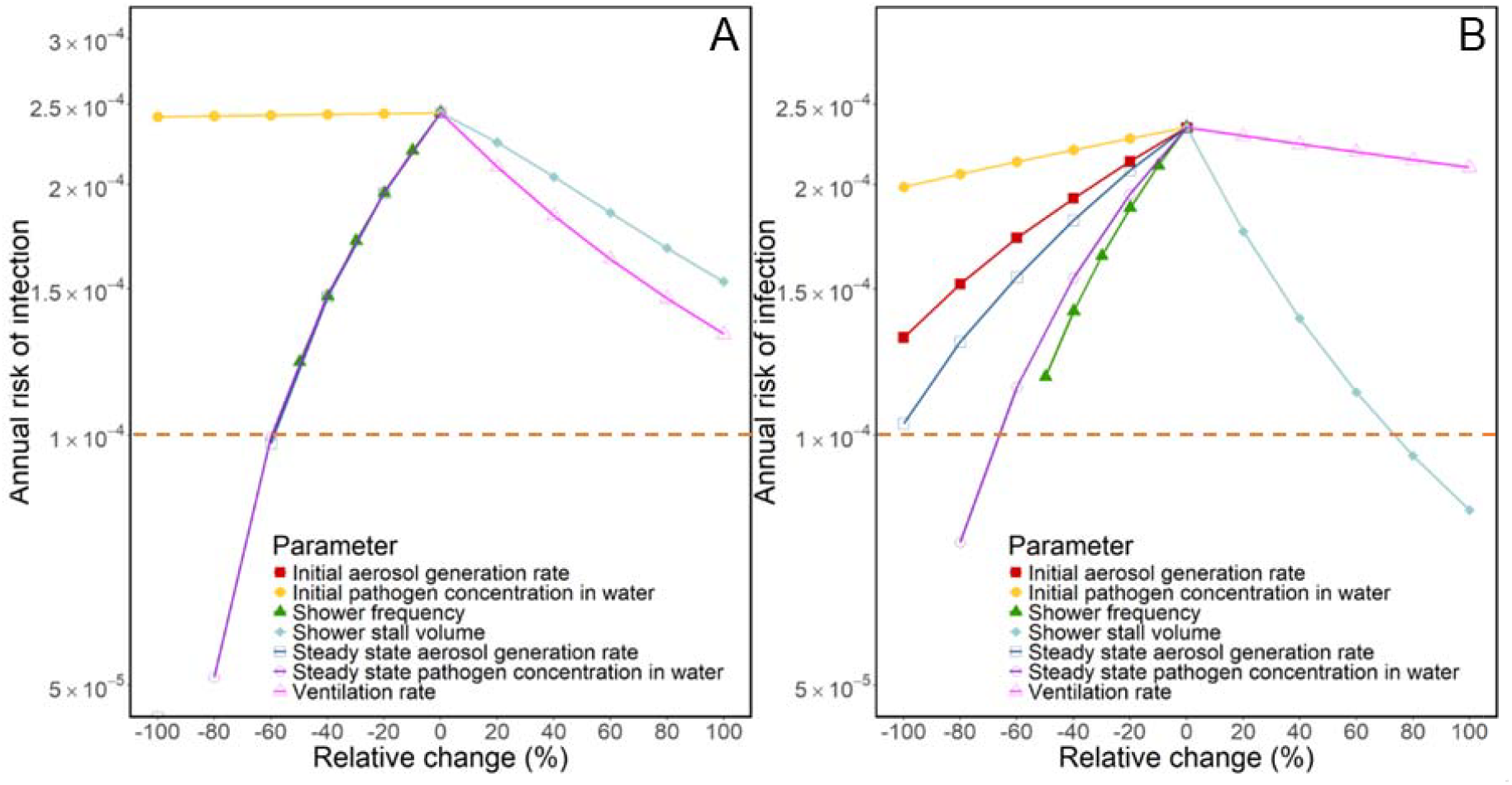
Alternative scenario modeling for cold showers (A) and hot showers (B) using a conventional showerhead. Relative change refers to the percentage change of parameters from the baseline values. The red dashed line represents the infection risk benchmark of 10^-4^ infections pppy.

## 4. Discussion

Application of a QMRA framework to calculate the risk of infection from exposure to *L. pneumophila* via showering that incorporated dynamic aerosol and pathogen concentrations highlighted elevated infection risks, including for adults and the elderly. The empirical *L. pneumophila* concentrations sampled in showers in the school in Switzerland indicated that the population is at risk for infection above both commonly accepted benchmark risks of 10^-4^ infections per person per year if taking hot showers. It should be noted that this school is not representative of all schools in Switzerland, but was identified for a sampling campaign due to previously known *Legionella* spp. contamination, and so we used it as an example of an at-risk building. For hot shower scenarios, the infection risks could exceed the benchmark value within the first few minutes of showers. Incorporating dynamic aerosol and pathogen concentrations suggested critical *L. pneumophila* concentrations in both first draw samples and flush samples should be considered for routine water monitoring especially at hot water temperatures.

### 4.1. Risk mitigation strategies and impacts on predicted risks

Our study confirms that control of *L. pneumophila* concentrations in building plumbing systems remains the most effective way of reducing risks in shower exposures.^48, 49^ Concentrations of *L. pneumophila* can be controlled by multiple strategies such as chemical disinfection, thermal inactivation, and regular flushing.^50-52^. Chemical disinfection is a commonly supported method for reducing *L. pneumophila* reductions in drinking water systems.^53, 54^ However, water stagnation and temperature can influence the decay of residual chlorine and therefore influence the effectiveness of chlorine disinfection.^29^ Although chlorination of plumbing systems is common in most countries like the US and China, in some countries (including Switzerland) drinking water is distributed without residual chlorine. Compared to maintaining residual chlorines at distal points, chlorine-free drinking water requires more careful protection of water sources, better treatment of drinking water prior to distribution, and sufficient management of distribution systems.

The effects of ventilation for risk reduction are influenced by site-specific conditions. In our specific scenario, we showed ventilation was not effective for risk reduction based on our alternative scenario modeling. Ventilation for showers is likely an important intervention in other contexts, for example small spaces with limited initial ventilation that otherwise allow the build-up of aerosol concentrations. Ventilation can also be a relatively simple intervention strategy, even if its effectiveness relative to other factors is low. Studies of respiratory pathogens such as MERS-CoV and SARS-CoV-2 highlight that ventilation is effective, but may be less effective than more intensive interventions such as wearing masks.^47, 55^ Risk reductions by ventilation may be more effective on intermittent emissions as it reduces the initial high concentrations of pathogens, whereas ventilation on continuous emissions reduces the steady state pathogen concentrations.^56^

Selection of appropriate showerheads is a potential way of reducing the risk of infections. Our modeling showed that reducing aerosol generation rates can have similar relative risk reductions as reducing *L. pneumophila* concentrations. Using water-saving showerheads with lower flow rates can potentially reduce the aerosol generation rates by 1-2 log_10_ compared to conventional showerheads.^2, 26, 57^ However, the performance of a showerhead may also change over time. Showerhead ages, for example, were shown to be more influential than water flow rates on microbial exposures as the showerhead ages were correlated with biofilm densities of opportunistic pathogens.^58^ Therefore, controlling the biofilm formation of showerheads, which acts to reduce the initial concentrations of *L. pneumophila*, may be more effective than applying water-saving showerheads.

### 4.2. Implications of predicted critical *L. pneumophila* concentrations on sampling strategies for regular water monitoring

By implementing our model, we were able to estimate critical concentrations (i.e., concentrations in flush samples exclusive of the first liter) of *L. pneumophila* that lead to risks of infections equal to a risk benchmark of 10^-4^ infections pppy. Our model output suggests *L. pneumophila* concentrations should be kept below 1000 CFU/L for scenarios in which aerosol generation rates match the rates from our cold showers and 25 CFU/L for scenarios similar to our hot showers. These values aligned with Wilson, et al. ^59^, and were generally 1-2 log_10_ higher than values reported by other previous QMRA studies using the subclinical infection endpoint^2, 22^. The reason that we predicted less stringent critical *L. pneumophila* concentrations than previous QMRA studies is due to lower aerosol generation rates applied in our study.^26, 33^ For cold showers, we also considered a gradual increase of aerosol concentration during showers instead of using the steady state concentrations for the whole shower duration. This gradual increase lowers the estimated risk relative to the assumption of steady state conditions.

The predicted critical concentrations at which risks exceed a benchmark of 10^-4^ infections pppy in first draw samples were 2-3 log_10_ higher than the flush samples. The simulation of scenarios where steady state *L. pneumophila* concentrations were assumed to be uncontaminated (i.e., 0 CFU/L) suggested that initial high concentrations in first draw samples could lead to risks above the benchmark especially for hot showers. For showers with varying durations, the importance of steady state *L. pneumophila* concentrations relative to initial concentrations may vary, with first flush samples becoming more important for shorter showers. In most of the samples collected, we showed that the initial concentrations were higher than steady state concentrations. The observed higher initial concentrations compared to steady state concentrations during flushing were consistent with previous studies.^27, 30, 31^ This finding, if it is replicable at other sites, suggests that regular monitoring first draw sample concentrations at distal points can help determine the extent to which further sampling for different locations within the circulation loops is needed.

### 4.3. Limitations of dynamic QMRA framework

We note the following limitations for our developed QMRA framework. Our dynamic modeling of aerosol and *L. pneumophila* concentrations remain site-specific, as some of the model parameters (notably *L. pneumophila* concentrations and aerosol generation rates) were calibrated based on limited site-specific data. This finding highlights the need to characterize the studied system. Alternatively, furthering the mechanistic modeling of pathogen (e.g., microbial growth) and aerosol concentrations (e.g., computational fluid dynamics) may be beneficial to generalize QMRA frameworks to multiple scenarios. ^60, 61^ A second limitation is that the modeled *L. pneumophila* concentrations are based on empirical measurements. Our measurements and modeling of temporal changes in *L. pneumophila* are based on two samples: the first liter sample and an average concentration estimated for the remaining 4 L samples. To better validate the releasing profile of *L. pneumophila* during flushing, better time resolution of *L. pneumophila* concentrations is needed. For example, Grimard-Conea, Deshommes, Doré and Prévost ^27^ analyzed total cell counts in each 0.4 liter sample for the first minute flushing and produced a complete bacteria releasing profile. Finally, the survival of *L. pneumophila* in aerosols can be affected by environmental conditions such as temperature and relative humidity.^62, 63^ Previous studies have suggested that viable *L. pneumophila* in aerosols only account for small fraction of *L. pneumophila* in water. ^64^ For waterborne pathogens such as *Brevundimonas diminuta* and *Pseudomonas aeruginosa*, the hot water conditions can reduce the culturability of bioaerosols.^65^ Including viability and associated risks of *L. pneumophila* in QMRA models may give better risk estimates for different environmental conditions. In addition to viability, the infectivity is also uncertain as current dose-response models mainly focus on *L. pneumophila* serogroup 1.^18^ The dose-response parameter was also identified as one of the most influential parameters in our sensitivity analyses, and further study on infectivity of *L. pneumophila* strain type combined with strain typing of environmental samples would help to address uncertainty of scenario-specific risk estimates.

## 5. Conclusion

Our developed QMRA indicated that annual infection risks above a typically acceptable risk benchmark are reached within the first minute of hot showers for the modeled *L. pneumophila* concentrations, which were observed in a building in Switzerland at known risk for legionellosis cases. Within this building, we observed positive correlations between the first draw sample concentrations and the flush sample concentrations. In accounting for this relationship, our predicted critical concentrations for flush samples that correspond to risks below the benchmark risk of 10^-4^ infections per person per year, are approximately consistent with current *L. pneumophila* concentration guidelines. Based on the results of a sensitivity analysis, controlling *L. pneumophila* concentrations in plumbing systems is the most effective risk mitigation strategy. Within routine monitoring programs, first draw samples and flush samples should be both considered to inform potential risks. In addition to providing practical insights on risks from *L. pneumophila* in showers, the study offers a framework for incorporating impacts of dynamic aerosol and pathogen concentrations into microbial risk assessment models.

## 6. Acknowledgements

We gratefully acknowledge financial support from the Swiss Federal Food Safety and Veterinary Office (FSVO), the Federal Office of Public Health (FOPH) and the Federal Office of Energy (FOE), through the project LeCo (Legionella Control of Buildings; Aramis nr.: 4.20.01).

## 7. Data Availability Statement

The data for the study are available at: https://doi.org/10.25678/000FNM. The code is available at: https://github.com/EawagPHH/QMRA_Legionella.

## 8. Author Contributions (CRediT Statement)

**LT:** Conceptualization, Methodology, Investigation, Formal Analysis, Data Curation, Software, Visualization, Validation, Writing – Original Draft, Writing – Review and Editing. **ES:** Conceptualization, Methodology, Software, Writing – Review and Editing. **KAH:** Conceptualization, Writing – Review and Editing. **FH:** Conceptualization, Resources, Writing – Review and Editing, Project administration, Funding acquisition. **TRJ:** Conceptualization, Resources, Supervision, Writing – Review and Editing, Project administration, Funding acquisition.

