## Supplemental Information for "Impacts of dynamic aerosol and pathogen concentrations on risks of *Legionella pneumophila* for Public Showers in Switzerland Based on a Quantitative Microbial Risk Assessment Framework"

**Text S1. Dynamic change of aerosol concentration in shower stall**

Previous studies have reported that aerosol concentrations in showers increase exponentially until reaching steady state.^1, 2^ Based on this, we applied a mass balance model to describe the fate and transport of generated aerosols assuming well-mixed conditions within the shower stall. The mass balance model is described by equation (1):

$V_{m}\frac{dC_{aer,i}}{dt}=-\left( Q+\sum v_{i,j}S_{j}+\lambda_{i}V_{m} \right)\cdot C_{aer,i}+G_{i}$	(1)

where *V_m_* is the volume of the shower stall (m^3^); *C_aer_,_i_* is the mass concentration of aerosol of size class i assuming the aerosols are pure water droplets (g/m^3^); *t* is the exposure duration (min); *Q* is the ventilation rate or air flow rate (m^3^/min); *S_j_* is the area of vertical, upward-facing, and downward-facing surface j within the shower stall (m^2^); *v_i,j_* is the deposition velocity of aerosol of aerosol of size i for the surface j (m/min); *λ_i_* is the aerosol residual decay rate caused by other removal process excluding deposition and ventilation rate for aerosol of size i (1/min); *G* is the generation rate of aerosol of size class i (g/min). The deposition velocities were calculated following the method proposed by Lai and Nazaroff ^3^. Other parameter values including aerosol generation rates, aerosol decay rates, and ventilation rates can be accessed in supporting information (Table S1).

Text S2. Dynamic change of concentrations of *L. pneumophila* during flushing

In our study, we observed that the first draw samples, which represented the 1^st^ liter collected, contained higher concentrations of *Legionella* spp. than the flush samples, which represented the 5^th^ liters collected. For our risk assessment model, we assume the composite flush sample concentrations represent the average concentrations in circulation loops of plumbing systems. We also assume that the *L. pneumophila* were evenly distributed in the circulation loops and only bulk water concentrations were considered (i.e., biofilm detachment process was negligible). Therefore, the dynamic change of concentrations of *L. pneumophila* was modeled as:

$\frac{dC_{leg}(t)}{dt}=\frac{Q_{w}}{V_{w}}(C_{leg,cir}-C_{leg}\left( t \right))$ (2)

Numerically solving equation (2) yields equation (3):

$C_{leg}\left( t \right)=C_{leg,cir}-(C_{leg,cir}-C_{leg}\left( 0 \right))e^{-\frac{Q_{w}}{V_{w}}\cdot t}$ (3)

Where *t* is the shower duration (min); *Q_w_* is the water flow rate (m^3^/min); *V_w_* is the volume of the first flush from the water pipes connecting to the showerheads (1L, or 0.001 m^3^ in this study); *C_leg,cir_* is the concentration of *L. pneumophila* in circulation loops (CFU/L); *C_leg_(t)* is the concentration of *L. pneumophila* at each time point (CFU/L).

For the initial condition, we assumed our measured first draw sample concentrations represented the initial contamination within the pipe at time t = 0, or *C_leg_(0)*. We assumed our flush sample concentrations represented the concentrations in the circulation loops, or *C_leg,cir_*. As this is a conservative assumption that may lead to overestimated risks, we also simulated scenarios where steady state concentrations of source water were assumed to be uncontaminated, equivalent to 0 CFU/L.

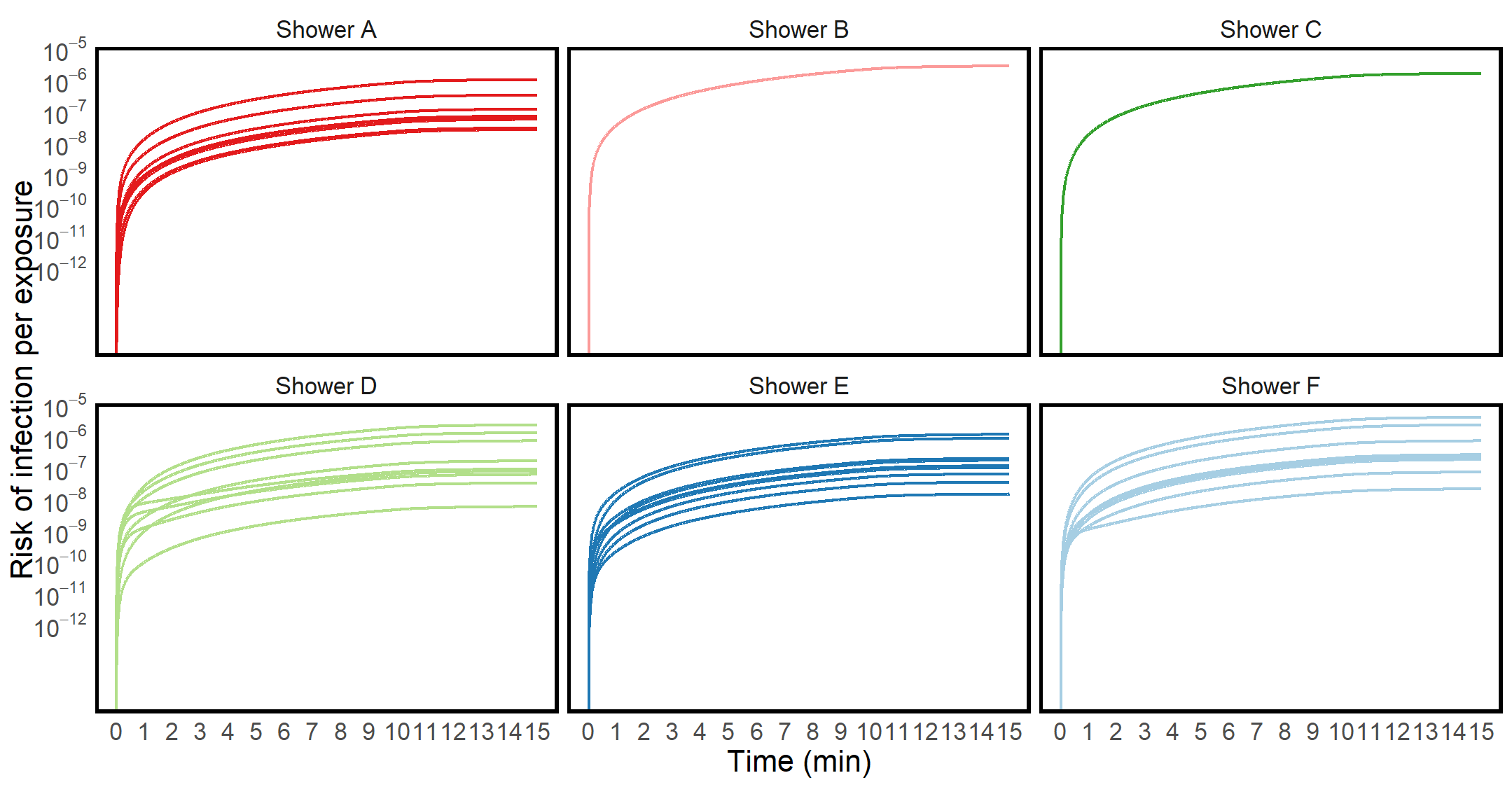

Figure S1. Median cumulative risks per exposure for each shower at cold showers. The lines in the same panel represent risks on different sampling dates.

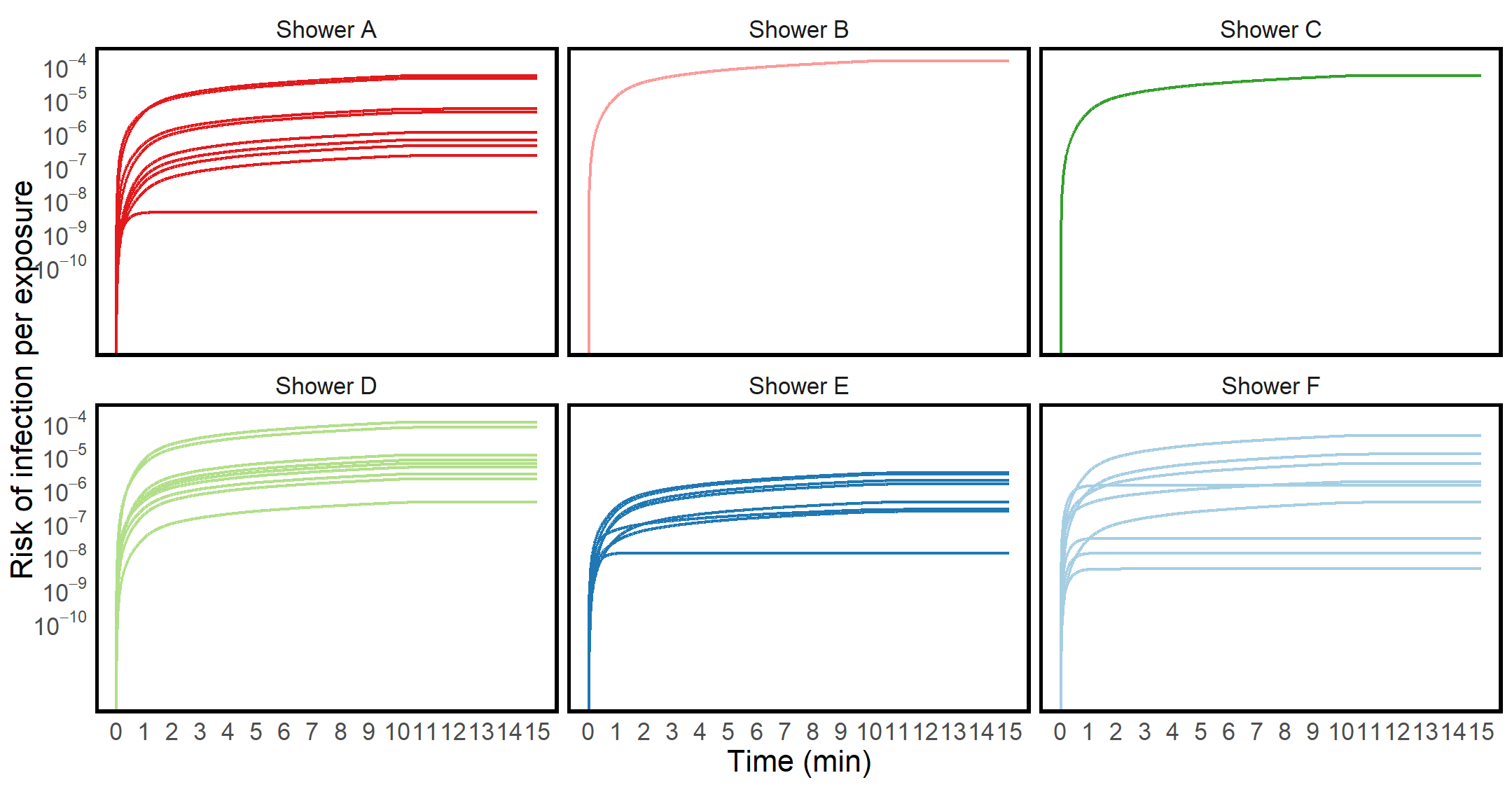

Figure S2. Median cumulative risks per exposure for each shower at hot showers. The lines in the same panel represent risks on different sampling dates.

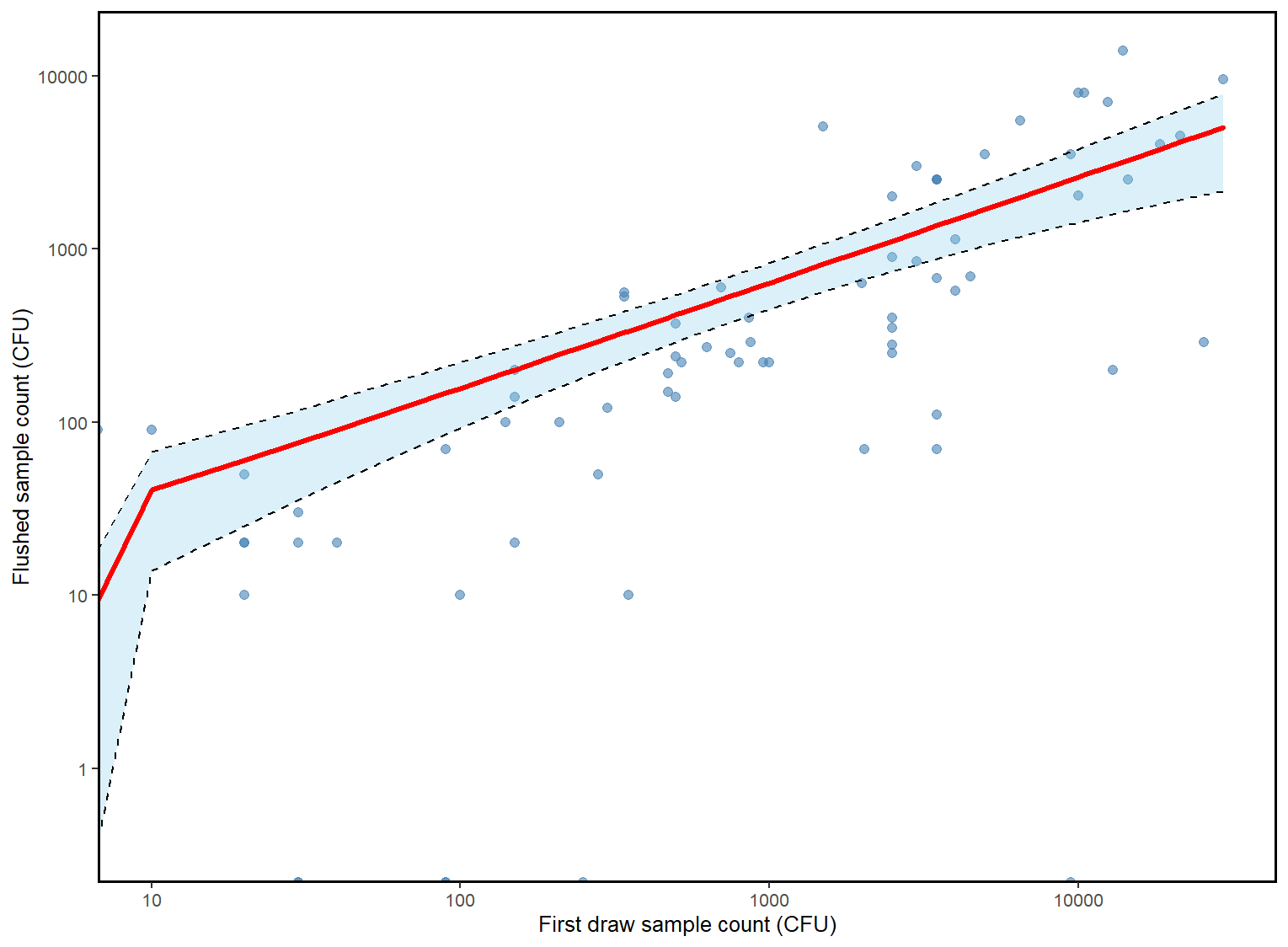

Figure S3. Associations between concentrations of *L. pneumophila* in first draw samples and flushed samples based on negative-binomial regression model.

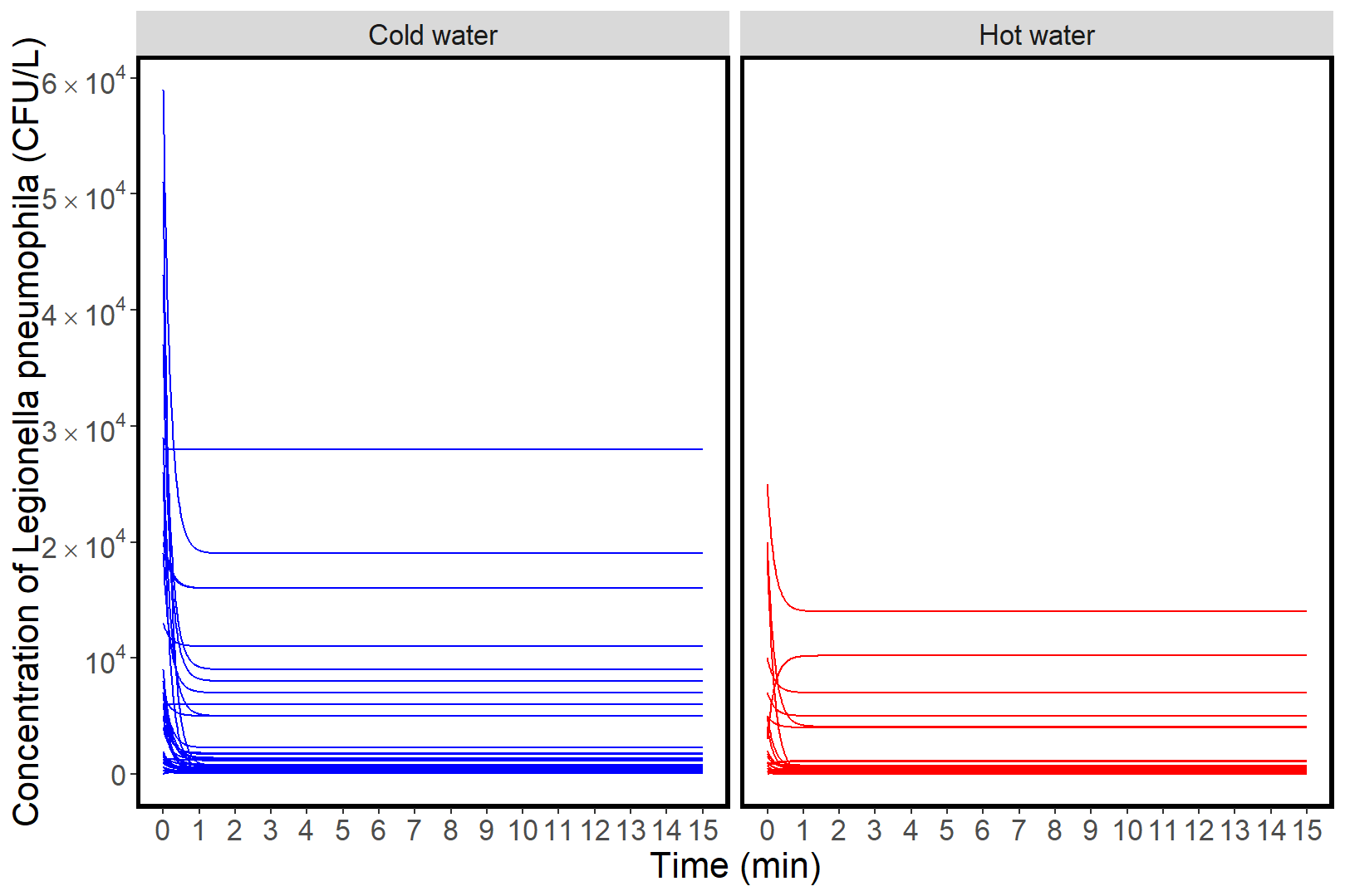

Figure S4. Dynamic concentrations of *L. pneumophila* simulated based on first and fifth liter samples.

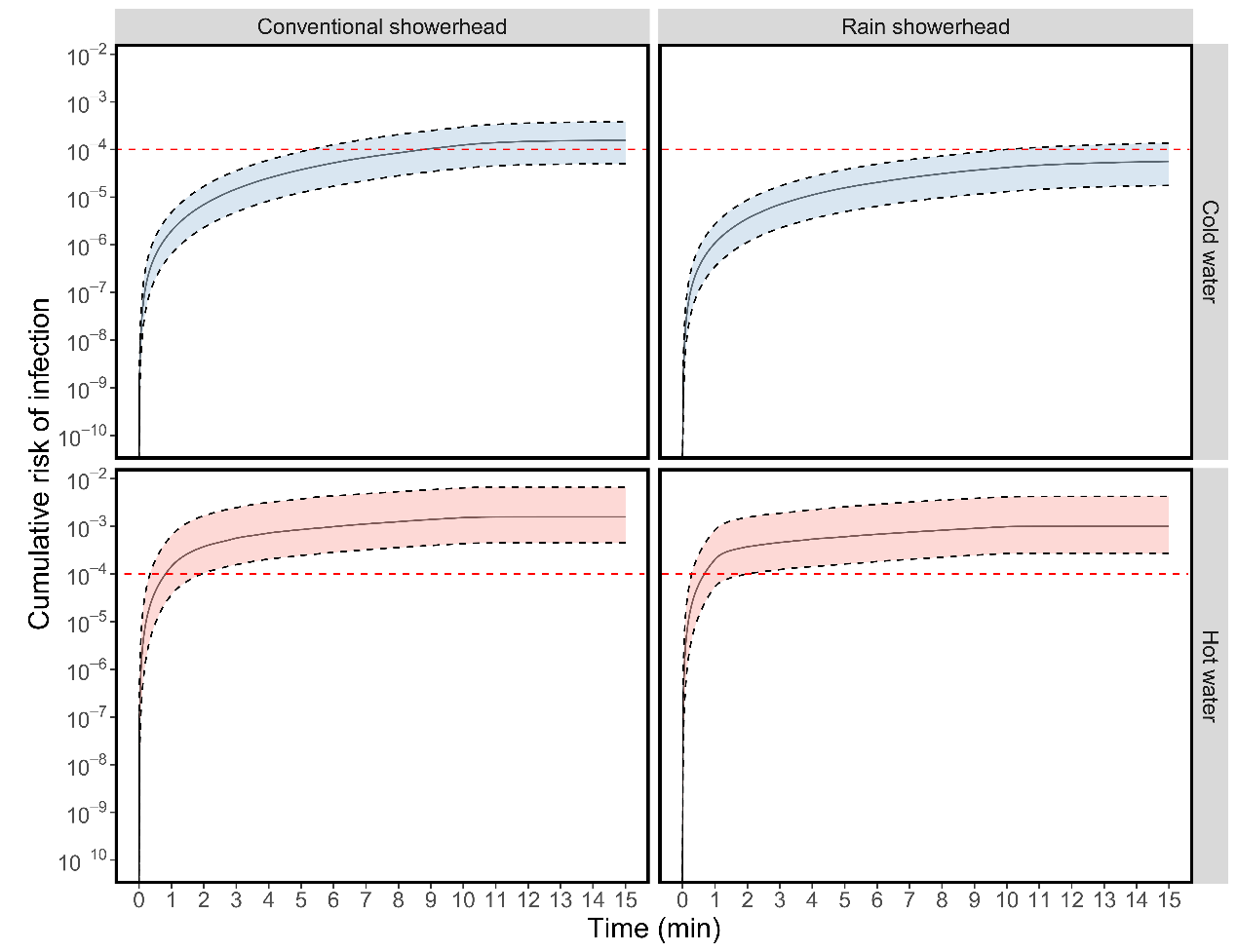

Figure S5. Cumulative annual risks of infection during a typical 15-minute shower based on empirical measurements from a public shower in Switzerland assuming similar concentration profiles during each exposure event. Solid black lines represent median risks. Black dashed lines represent 25% percentile and 75% percentile values of predicted risks. Shaded areas represent the 50% uncertainty intervals. Red dashed horizontal lines reflect the risk threshold of 10^-4^ infections per year.

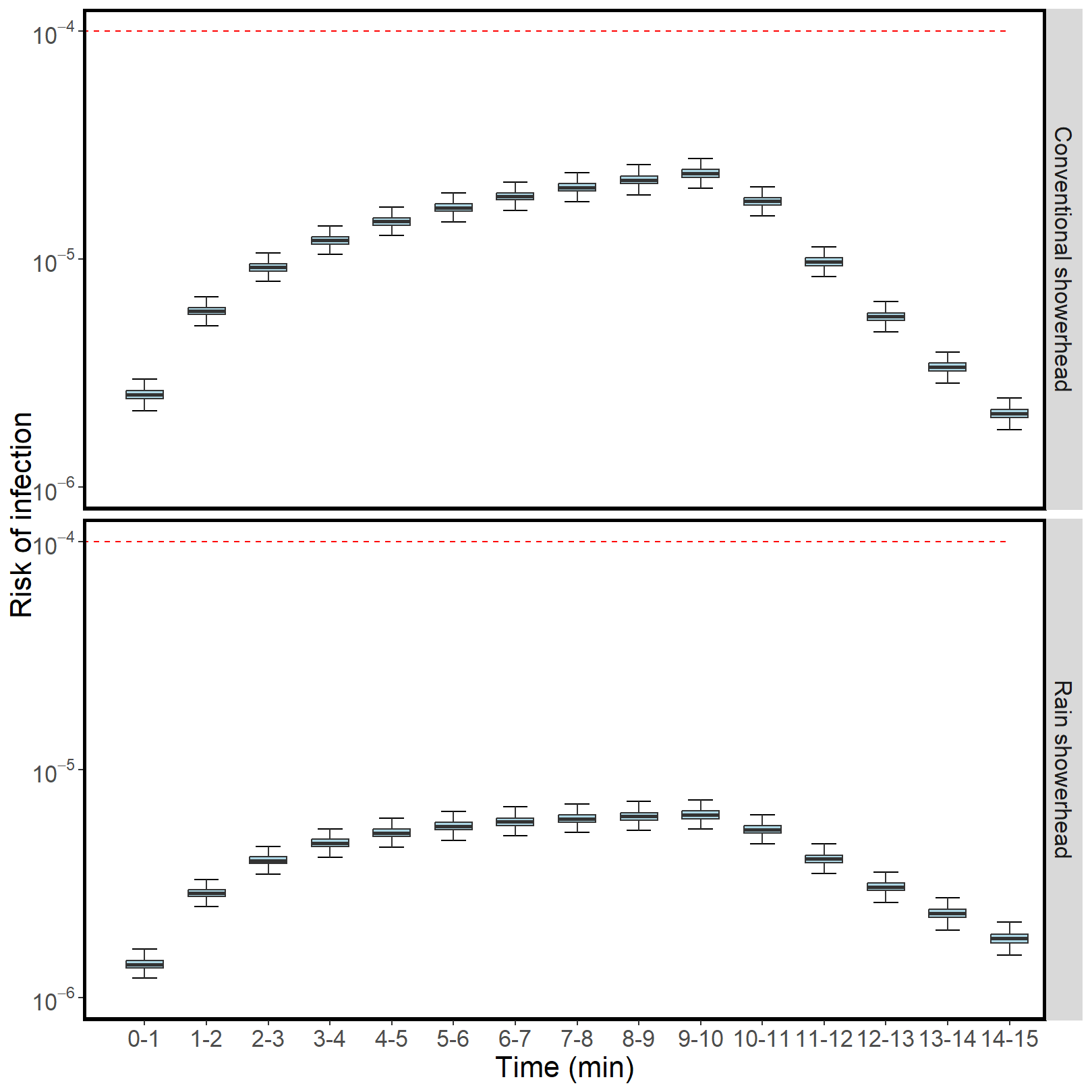

Figure S6. The risk of infection (annualized) for discrete time periods during a typical 15-minute cold shower. Boxplots represent the median and interquartile range, with whiskers representing the minimum to maximum range. Red horizontal dashed lines represent the risk threshold of 10^-4^ infections per year.

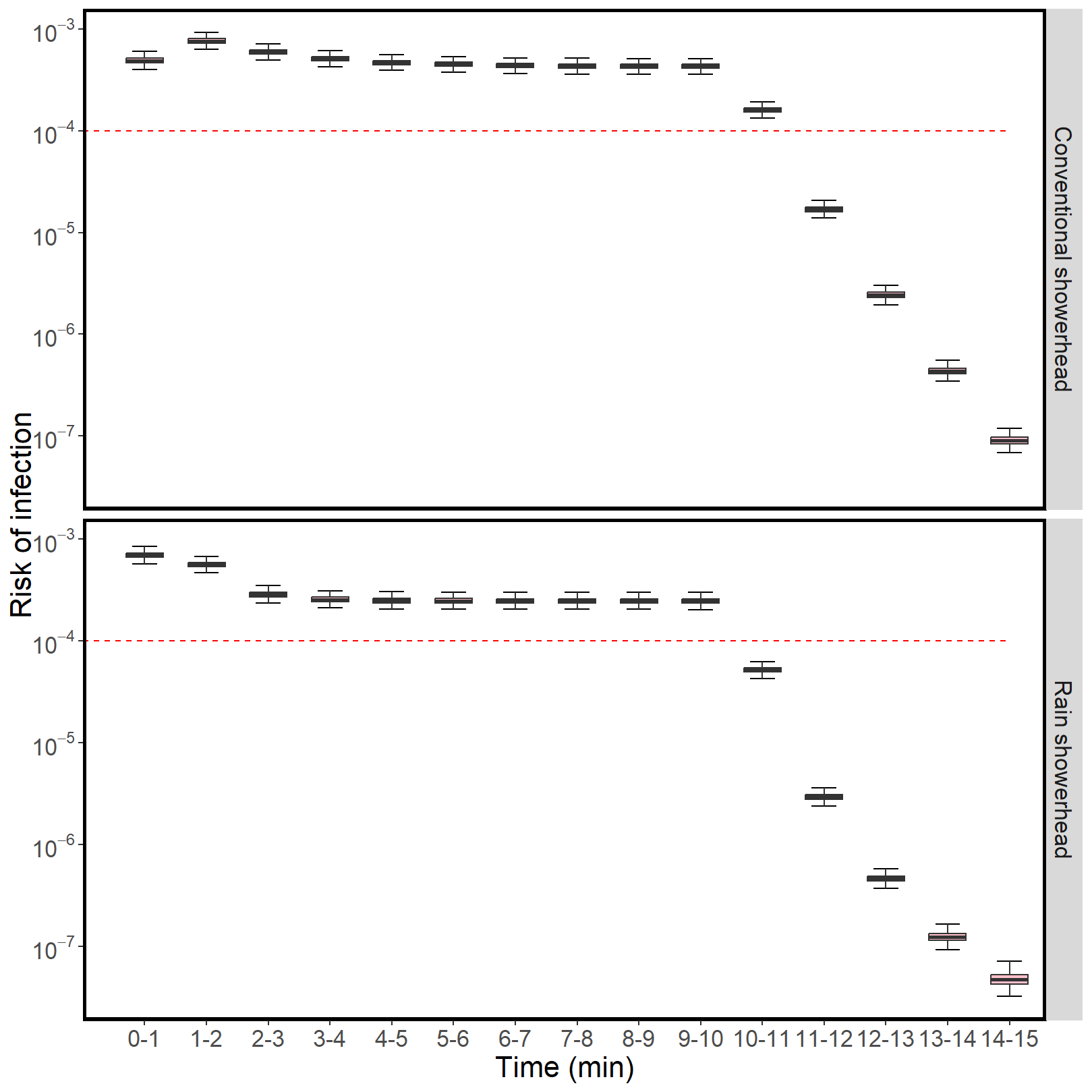

Figure S7. The risk of infection (annualized) for discrete time periods during a typical 15-minute hot shower. Boxplots represent the median and interquartile range, with whiskers representing the minimum to maximum range. Red horizontal dashed lines represent the risk threshold of 10^-4^ infections per year.

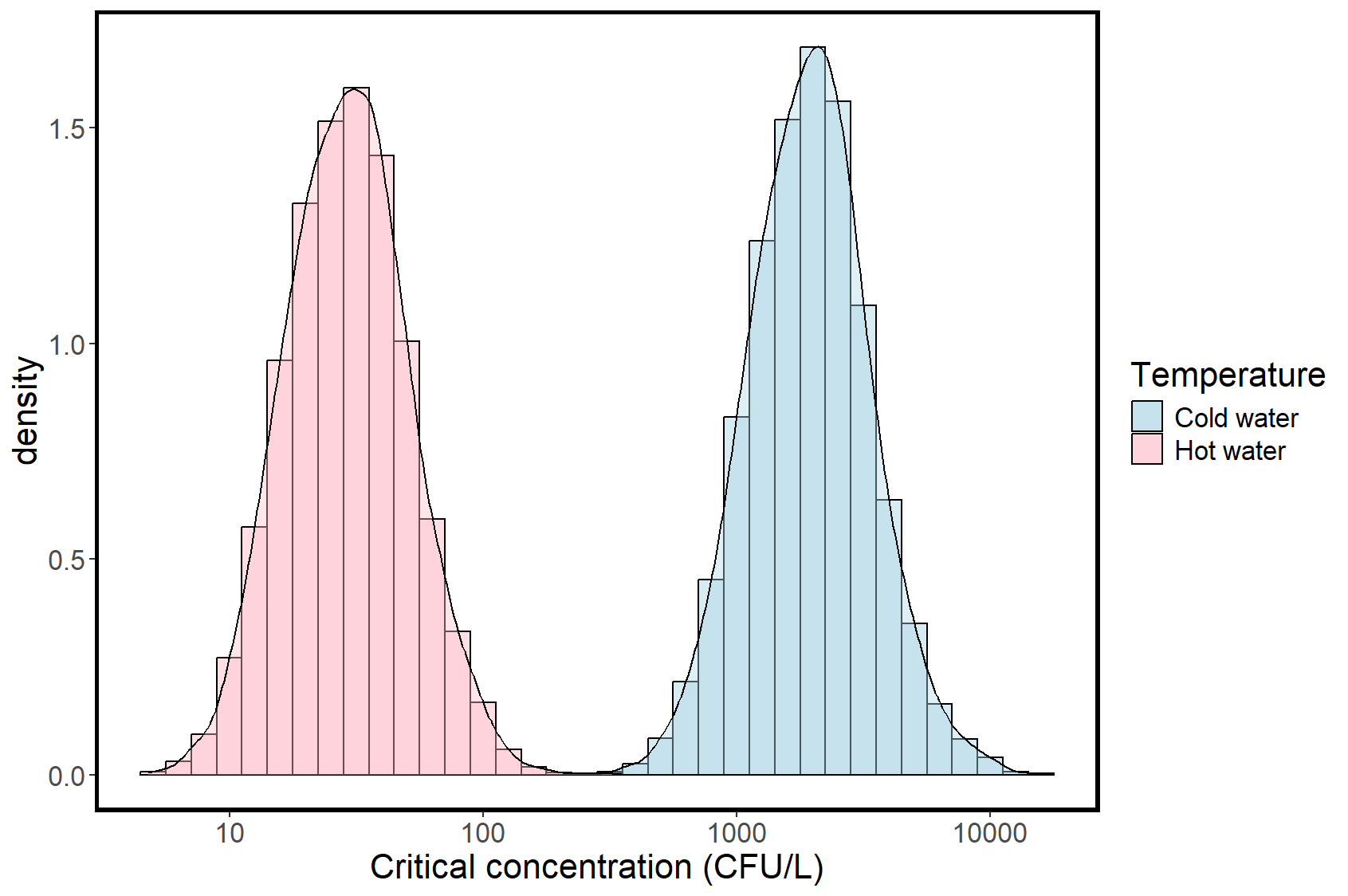

Figure S8. Probability density function (PDF) of model estimated critical *L. pneumophila* concentrations leading to a risk threshold of 10^-4^ infections per year.

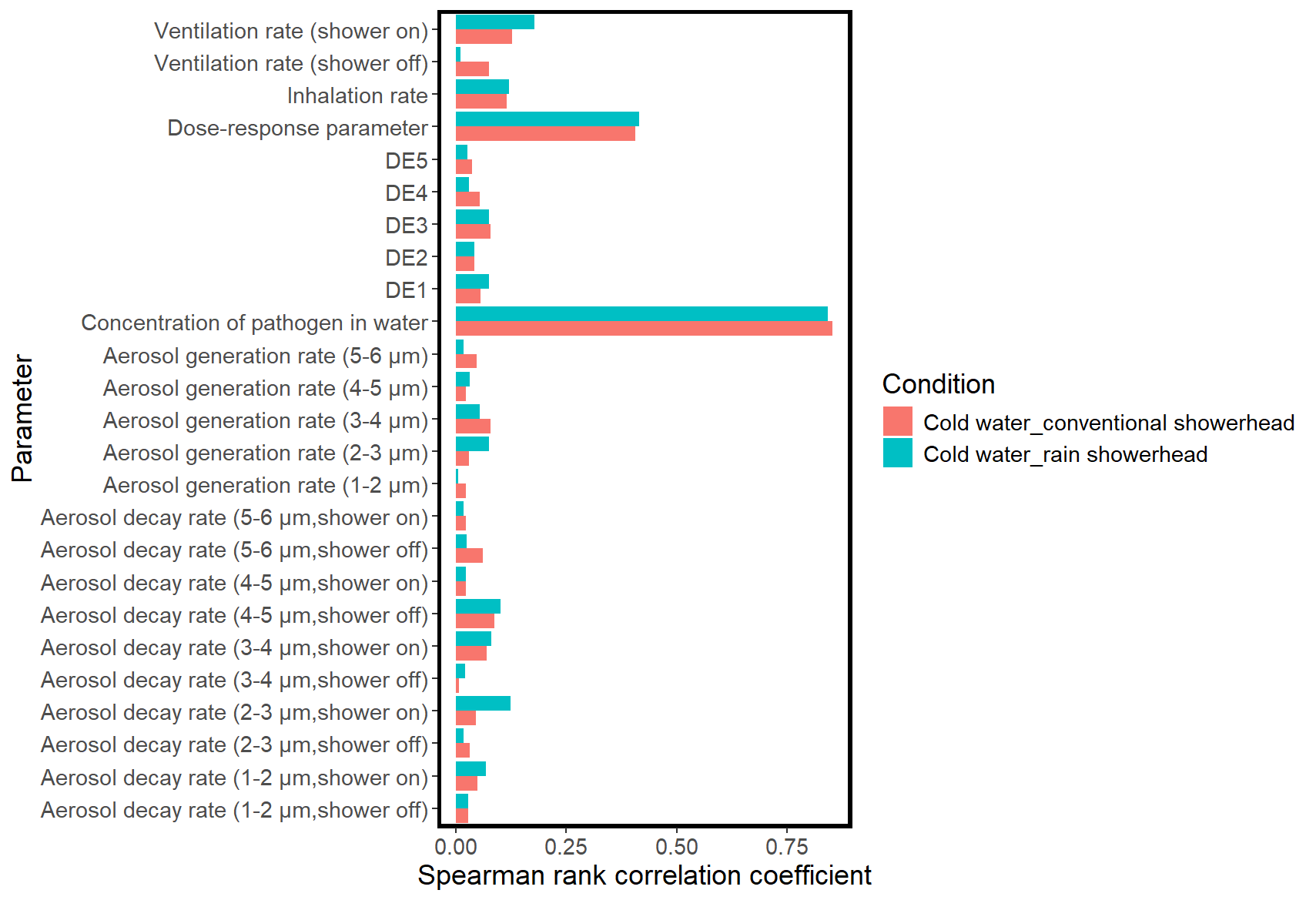

Figure S9. Sensitivity analysis for cold shower scenarios showing the magnitude of the Spearman rank correlation coefficient between the variation in parameter values and the estimated infection risk. DE refers to deposition efficiency of aerosols at alveoli.

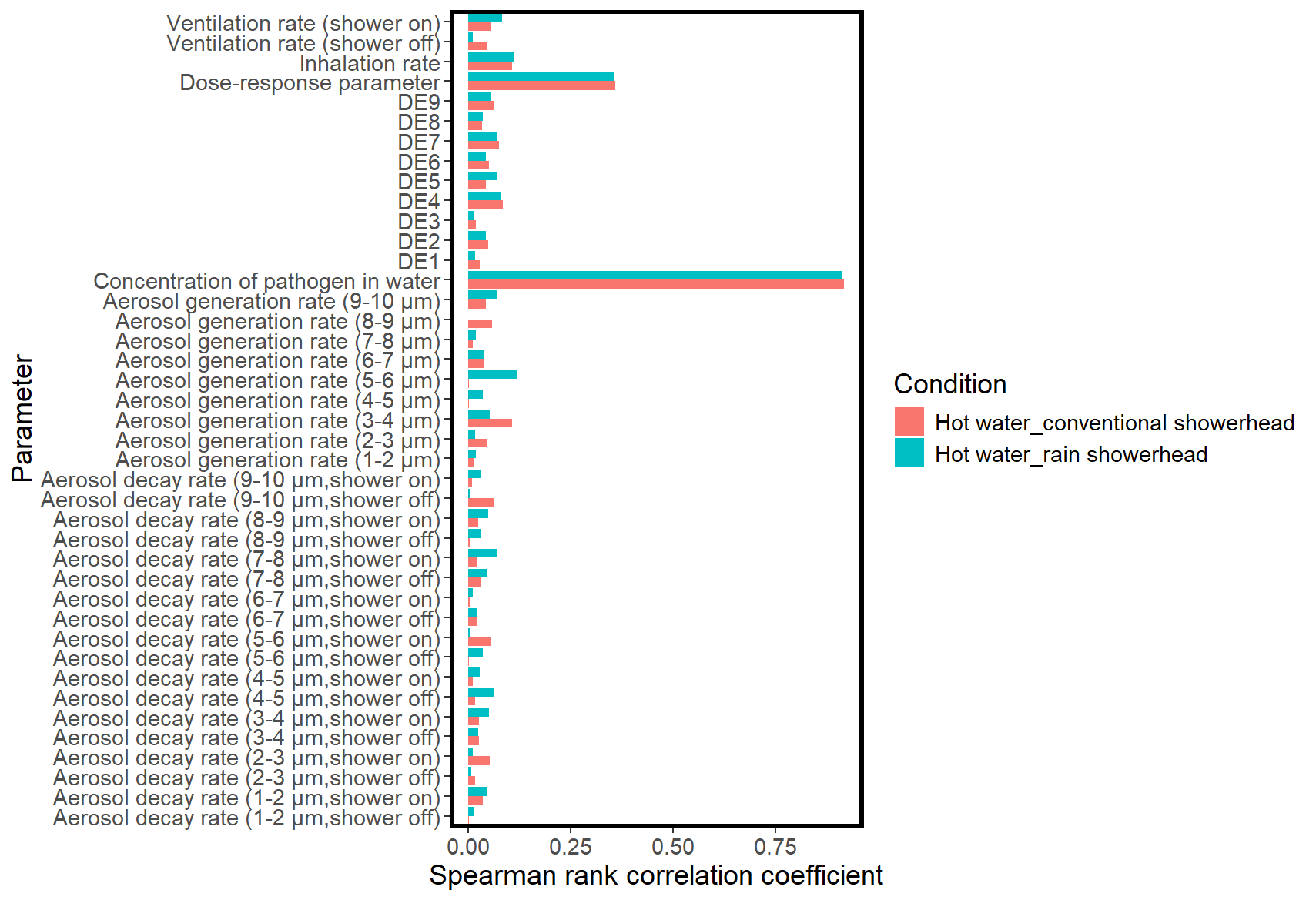

Figure S10. Sensitivity analysis for hot shower scenarios showing the magnitude of the Spearman rank correlation coefficient between the variation in parameter values and the estimated infection risk. DE refers to deposition efficiency of aerosols at alveoli.

**Table S1**. Monte Carlo input model parameters

| Parameter | Symbol | Distribution | Value | Unit | Source |
| --- | --- | --- | --- | --- | --- |
| Aerosol generation rate | G_i_ |  |  |  |  |
| Cold water |  |  |  |  |  |
| Conventional showerhead |  |  |  |  |  |
| 1-2 μm |  | Uniform | min=2.0e-2, max=2.3e-2 | mg/min | Tang, et al. ^4^ |
| 2-3 μm |  | Uniform | min=2.7e-2, max=4.4e-2 | mg/min | Tang, et al. ^4^ |
| 3-4 μm |  | Uniform | min=3.1e-2, max=8.7e-2 | mg/min | Tang, et al. ^4^ |
| 4-5 μm |  | Uniform | min=2.5e-2, max=6.8e-2 | mg/min | Tang, et al. ^4^ |
| 5-6 μm |  | Uniform | min=2.0e-2, max=1.2e-1 | mg/min | Tang, et al. ^4^ |
| Rain showerhead |  |  |  |  | Tang, et al. ^4^ |
| 1-2 μm |  | Uniform | min=1.7e-2, max=2.2e-2 | mg/min | Tang, et al. ^4^ |
| 2-3 μm |  | Uniform | min=2.2e-2, max=3.0e-2 | mg/min | Tang, et al. ^4^ |
| 3-4 μm |  | Uniform | min=2.3e-2, max=3.5e-2 | mg/min | Tang, et al. ^4^ |
| 4-5 μm |  | Uniform | min=1.5e-2, max=2.7e-2 | mg/min | Tang, et al. ^4^ |
| 5-6 μm |  | Uniform | min=7e-3, max=3.1e-2 | mg/min | Tang, et al. ^4^ |
| Hot water |  |  |  |  |  |
| Conventional showerhead |  |  |  |  |  |
| The first minute |  |  |  |  |  |
| 1-2 μm |  | Uniform | min=2.2e-1, max=1.0 | mg/min | Tang, et al. ^4^ |
| 2-3 μm |  | Uniform | min=2.0e-1, max=6.6 | mg/min | Tang, et al. ^4^ |
| 3-4 μm |  | Uniform | min=4.1e-1, max=3.1e1 | mg/min | Tang, et al. ^4^ |
| 4-5 μm |  | Uniform | min=6.2e-1, max=7.6e1 | mg/min | Tang, et al. ^4^ |
| 5-6 μm |  | Uniform | min=7.9e-1, max=2.1e2 | mg/min | Tang, et al. ^4^ |
| 6-7 μm |  | Uniform | min=1.4e-1, max=2.1e2 | mg/min | Tang, et al. ^4^ |
| 7-8 μm |  | Uniform | min=1.1e-1, max=1.9e2 | mg/min | Tang, et al. ^4^ |
| 8-9 μm |  | Uniform | min=1.2e-1, max=1.5e2 | mg/min | Tang, et al. ^4^ |
| 9-10 μm |  | Uniform | min=7.2e-2, max=1.7e2 | mg/min | Tang, et al. ^4^ |
| After the first minute |  |  |  |  |  |
| 1-2 μm |  | Uniform | min=3.4e-2, max=5.6e-1 | mg/min | Tang, et al. ^4^ |
| 2-3 μm |  | Uniform | min=5.3e-2, max=2.3 | mg/min | Tang, et al. ^4^ |
| 3-4 μm |  | Uniform | min=8.0e-2, max=9.4 | mg/min | Tang, et al. ^4^ |
| 4-5 μm |  | Uniform | min=1.0e-1, max=1.8e1 | mg/min | Tang, et al. ^4^ |
| 5-6 μm |  | Uniform | min=9.5e-2, max=7.5e1 | mg/min | Tang, et al. ^4^ |
| 6-7 μm |  | Uniform | min=2.4e-2, max=30.3 | mg/min | Tang, et al. ^4^ |
| 7-8 μm |  | Uniform | min=1.1, max=5.3e1 | mg/min | Tang, et al. ^4^ |
| 8-9 μm |  | Uniform | min=7.2e-3, max=7.2e1 | mg/min | Tang, et al. ^4^ |
| 9-10 μm |  | Uniform | min=4.6e-3, max=5.0e1 | mg/min | Tang, et al. ^4^ |
| Rain showerhead |  |  |  |  |  |
| The first minute |  |  |  |  |  |
| 1-2 μm |  | Uniform | min=1.7e-1, max=1.32 | mg/min | Tang, et al. ^4^ |
| 2-3 μm |  | Uniform | min=9.5e-1, max=9.1 | mg/min | Tang, et al. ^4^ |
| 3-4 μm |  | Uniform | min=3.2, max=5.0e1 | mg/min | Tang, et al. ^4^ |
| 4-5 μm |  | Uniform | min=8.2, max=1.4e2 | mg/min | Tang, et al. ^4^ |
| 5-6 μm |  | Uniform | min=14.6, max=3.2e2 | mg/min | Tang, et al. ^4^ |
| 6-7 μm |  | Uniform | min=11.0, max=1.7e2 | mg/min | Tang, et al. ^4^ |
| 7-8 μm |  | Uniform | min=17.7, max=1.5e2 | mg/min | Tang, et al. ^4^ |
| 8-9 μm |  | Uniform | min=29.2, max=1.7e2 | mg/min | Tang, et al. ^4^ |
| 9-10 μm |  | Uniform | min=31.4, max=1.7e2 | mg/min | Tang, et al. ^4^ |
| After the first minute |  |  |  |  |  |
| 1-2 μm |  | Uniform | min=2.6e-2, max=5.8e-1 | mg/min | Tang, et al. ^4^ |
| 2-3 μm |  | Uniform | min=1.5e-1, max=4.7 | mg/min | Tang, et al. ^4^ |
| 3-4 μm |  | Uniform | min=8.2e-1, max=2.3e1 | mg/min | Tang, et al. ^4^ |
| 4-5 μm |  | Uniform | min=3.0, max=3.6e1 | mg/min | Tang, et al. ^4^ |
| 5-6 μm |  | Uniform | min=9.8, max=1.9e2 | mg/min | Tang, et al. ^4^ |
| 6-7 μm |  | Uniform | min=8.9, max=1.9e-1 | mg/min | Tang, et al. ^4^ |
| 7-8 μm |  | Uniform | min=1.7e1, max=5.0e1 | mg/min | Tang, et al. ^4^ |
| 8-9 μm |  | Uniform | min=2.1e1, max=8.3e1 | mg/min | Tang, et al. ^4^ |
| 9-10 μm |  | Uniform | min=2.4e1, max=5.5e1 | mg/min | Tang, et al. ^4^ |
| Aerosol residual decay rate | λ_i_ |  |  |  |  |
| Cold water |  |  |  |  |  |
| Conventional showerhead |  |  |  |  |  |
| During shower |  |  |  |  |  |
| 1-2 μm |  | Uniform | min=-3.4e-2, max=1.5e-1 | 1/min | Tang, et al. ^4^ |
| 2-3 μm |  | Uniform | min=-9.0e-3, max=7.0e-3 | 1/min | Tang, et al. ^4^ |
| 3-4 μm |  | Uniform | min=-2.3e-2, max=2.3e-2 | 1/min | Tang, et al. ^4^ |
| 4-5 μm |  | Uniform | min=-2.7e-1, max=3.9e-2 | 1/min | Tang, et al. ^4^ |
| 5-6 μm |  | Uniform | min=-2.1e-1, max=6.5e-2 | 1/min | Tang, et al. ^4^ |
| After shower |  |  |  |  |  |
| 1-2 μm |  | Uniform | min=3.3e-2, max=4.5e-1 | 1/min | Tang, et al. ^4^ |
| 2-3 μm |  | Uniform | min=1.2e-1, max=7.2e-1 | 1/min | Tang, et al. ^4^ |
| 3-4 μm |  | Uniform | min=2.0e-1, max=9.1e-1 | 1/min | Tang, et al. ^4^ |
| 4-5 μm |  | Uniform | min=2.1e-1, max=1.1 | 1/min | Tang, et al. ^4^ |
| 5-6 μm |  | Uniform | min=2.9e-1, max=1.2 | 1/min | Tang, et al. ^4^ |
| Rain showerhead |  |  |  |  |  |
| During shower |  |  |  |  |  |
| 1-2 μm |  | Uniform | min=-1.0e-1, max=1.7e-1 | 1/min | Tang, et al. ^4^ |
| 2-3 μm |  | Uniform | min=-1.8e-1, max=1.3e-1 | 1/min | Tang, et al. ^4^ |
| 3-4 μm |  | Uniform | min=-2.1e-1, max=1.6e-1 | 1/min | Tang, et al. ^4^ |
| 4-5 μm |  | Uniform | min=-2.0e-1, max=2.8e-1 | 1/min | Tang, et al. ^4^ |
| 5-6 μm |  | Uniform | min=-1.9e-1, max=1.1 | 1/min | Tang, et al. ^4^ |
| After shower |  |  |  |  |  |
| 1-2 μm |  | Uniform | min=-5.3e-2, max=2.9e-1 | 1/min | Tang, et al. ^4^ |
| 2-3 μm |  | Uniform | min=3.1e-2, max=3.4e-1 | 1/min | Tang, et al. ^4^ |
| 3-4 μm |  | Uniform | min=2.4e-2, max=3.2e-1 | 1/min | Tang, et al. ^4^ |
| 4-5 μm |  | Uniform | min=-3.1e-2, max=4.0e-1 | 1/min | Tang, et al. ^4^ |
| 5-6 μm |  | Uniform | min=-3.0e-2, max=3.0e-1 | 1/min | Tang, et al. ^4^ |
| Hot water |  |  |  |  |  |
| Conventional showerhead |  |  |  |  |  |
| During shower |  |  |  |  |  |
| 1-2 μm |  | Uniform | min=5.1e-1, max=1.2 | 1/min | Tang, et al. ^4^ |
| 2-3 μm |  | Uniform | min=4.7e-1, max=8.8e-1 | 1/min | Tang, et al. ^4^ |
| 3-4 μm |  | Uniform | min=4.7e-1, max=8.8e-1 | 1/min | Tang, et al. ^4^ |
| 4-5 μm |  | Uniform | min=4.0e-1, max=6.3e-1 | 1/min | Tang, et al. ^4^ |
| 5-6 μm |  | Uniform | min=4.7e-1, max=5.0e-1 | 1/min | Tang, et al. ^4^ |
| 6-7 μm |  | Uniform | min=1.7e-1, max=9.4e-1 | 1/min | Tang, et al. ^4^ |
| 7-8 μm |  | Uniform | min=3.4e-1, max=8.0e-1 | 1/min | Tang, et al. ^4^ |
| 8-9 μm |  | Uniform | min=7.4e-2, max=6.9e-1 | 1/min | Tang, et al. ^4^ |
| 9-10 μm |  | Uniform | min=-8.3e-3, max=4.2e-1 | 1/min | Tang, et al. ^4^ |
| After shower |  |  |  |  |  |
| 1-2 μm |  | Uniform | min=1.2, max=2.0 | 1/min | Tang, et al. ^4^ |
| 2-3 μm |  | Uniform | min=1.2, max=2.8 | 1/min | Tang, et al. ^4^ |
| 3-4 μm |  | Uniform | min=1.7, max=3.1 | 1/min | Tang, et al. ^4^ |
| 4-5 μm |  | Uniform | min=1.9, max=3.3 | 1/min | Tang, et al. ^4^ |
| 5-6 μm |  | Uniform | min=1.3, max=3.0 | 1/min | Tang, et al. ^4^ |
| 6-7 μm |  | Uniform | min=8.9e-1, max=3.1 | 1/min | Tang, et al. ^4^ |
| 7-8 μm |  | Uniform | min=8.6e-1, max=2.9 | 1/min | Tang, et al. ^4^ |
| 8-9 μm |  | Uniform | min=8.0e-1, max=2.8 | 1/min | Tang, et al. ^4^ |
| 9-10 μm |  | Uniform | min=6.8e-1, max=2.5 | 1/min | Tang, et al. ^4^ |
| Rain showerhead |  |  |  |  |  |
| During shower |  |  |  |  |  |
| 1-2 μm |  | Uniform | min=1.2, max=5.3 | 1/min | Tang, et al. ^4^ |
| 2-3 μm |  | Uniform | min=1.1, max=8.3 | 1/min | Tang, et al. ^4^ |
| 3-4 μm |  | Uniform | min=1.1, max=6.6 | 1/min | Tang, et al. ^4^ |
| 4-5 μm |  | Uniform | min=1.0, max=5.3 | 1/min | Tang, et al. ^4^ |
| 5-6 μm |  | Uniform | min=8.9e-1, max=3.2 | 1/min | Tang, et al. ^4^ |
| 6-7 μm |  | Uniform | min=1.6e-1, max=2.3 | 1/min | Tang, et al. ^4^ |
| 7-8 μm |  | Uniform | min=4.3e-1, max=2.2 | 1/min | Tang, et al. ^4^ |
| 8-9 μm |  | Uniform | min=5.5e-1, max=2.6 | 1/min | Tang, et al. ^4^ |
| 9-10 μm |  | Uniform | min=3.9e-1, max=2.2 | 1/min | Tang, et al. ^4^ |
| After shower |  |  |  |  |  |
| 1-2 μm |  | Uniform | min=2.2e-1, max=1.1 | 1/min | Tang, et al. ^4^ |
| 2-3 μm |  | Uniform | min=1.2, max=1.6 | 1/min | Tang, et al. ^4^ |
| 3-4 μm |  | Uniform | min=1.6, max=3.9 | 1/min | Tang, et al. ^4^ |
| 4-5 μm |  | Uniform | min=1.6, max=7.0 | 1/min | Tang, et al. ^4^ |
| 5-6 μm |  | Uniform | min=1.5, max=1.3e1 | 1/min | Tang, et al. ^4^ |
| 6-7 μm |  | Uniform | min=1.6, max=1.3e1 | 1/min | Tang, et al. ^4^ |
| 7-8 μm |  | Uniform | min=1.6, max=1.3e1 | 1/min | Tang, et al. ^4^ |
| 8-9 μm |  | Uniform | min=1.6, max=1.3e1 | 1/min | Tang, et al. ^4^ |
| 9-10 μm |  | Uniform | min=1.6, max=1.3e1 | 1/min | Tang, et al. ^4^ |
| Volume of shower stall | V_m_ | Point | 2.5 | m^3^ | Tang, et al. ^4^ |
| Ventilation rate | Q |  |  |  |  |
| Cold water conventional showerhead |  |  |  |  |  |
| Before shower |  | Uniform | min=8.5e-1, max=9.6e- | m^3^/min | Tang, et al. ^4^ |
| During shower |  | Uniform | min=2.4e-1, max=5.5e-1 | m^3^/min | Tang, et al. ^4^ |
| After shower |  | Uniform | min=1.1e-1, max=5.5e-1 | m^3^/min | Tang, et al. ^4^ |
| Cold water rain showerhead |  |  |  |  |  |
| Before shower |  | Uniform | min=8.8e-2, max=5.5e-1 | m^3^/min | Tang, et al. ^4^ |
| During shower |  | Uniform | min=6.8e-1, max=1.5 | m^3^/min | Tang, et al. ^4^ |
| After shower |  | Uniform | min=0.0, max=6.6e-1 | m^3^/min | Tang, et al. ^4^ |
| Hot water conventional showerhead |  |  |  |  |  |
| Before shower |  | Uniform | min=0.0, max=9.8e-1 | m^3^/min | Tang, et al. ^4^ |
| During shower |  | Uniform | min=2.5e-1, max=6.3e-1 | m^3^/min | Tang, et al. ^4^ |
| After shower |  | Uniform | min=6.2e-1, max=1.2 | m^3^/min | Tang, et al. ^4^ |
| Hot water rain showerhead |  |  |  |  |  |
| Before shower |  | Uniform | min=0.0, max=4.4e-1 | m^3^/min | Tang, et al. ^4^ |
| During shower |  | Uniform | min=3.7e-1, max=1.1 | m^3^/min | Tang, et al. ^4^ |
| After shower |  | Uniform | min=3.4e-2, max=1.1 | m^3^/min | Tang, et al. ^4^ |
| Inhalation rate | Br | Uniform | min=1.3e-2, max=1.7e-2 | m^3^/min | EPA ^5^ |
| Deposition fraction | D_i_ |  |  |  | Heyder, et al. ^6^ |
| 1-2 μm |  | Uniform | min=2.3e-1, max=5.3e-1 | Unitless |  |
| 2-3 μm |  | Uniform | min=3.6e-1, max=6.2e-1 | Unitless |  |
| 3-4 μm |  | Uniform | min=2.9e-1, max=6.2e-1 | Unitless |  |
| 4-5 μm |  | Uniform | min=1.9e-1, max=6.1e-1 | Unitless |  |
| 5-6 μm |  | Uniform | min=1.0e-1, max=5.2e-1 | Unitless |  |
| 6-7 μm |  | Uniform | min=3.0e-2, max=4.0e-1 | Unitless |  |
| 7-8 μm |  | Uniform | min=3.0e-2, max=2.9e-1 | Unitless |  |
| 8-9 μm |  | Uniform | min=1.0e-2, max=1.9e-1 | Unitless |  |
| 9-10 μm |  | Uniform | min=1.0e-2, max=1.2e-1 | Unitless |  |
| Concentration of *L. pneumophila* | C_leg_ |  |  |  | Measured |
| Cold water |  | Poisson-gamma | shape=0.7, rate=5.9e-5 | CFU/L |  |
| Hot water |  | Poisson-gamma | shape=0.361, rate=1.1e-4 | CFU/L |  |
| *L. pneumophila* partitioning fraction | F_i_ |  |  |  | Allegra, et al. ^7^ |
| 1-2 μm |  | Point | 0.175 | Unitless |  |
| 2-3 μm |  | Point | 0.1639 | Unitless |  |
| 3-4 μm |  | Point | 0.1556 | Unitless |  |
| 4-5 μm |  | Point | 0.0667 | Unitless |  |
| 5-6 μm |  | Point | 0.0389 | Unitless |  |
| 6-7 μm |  | Point | 0.025 | Unitless |  |
| 7-8 μm |  | Point | 0.0278 | Unitless |  |
| 8-9 μm |  | Point | 0.05 | Unitless |  |
| 9-10 μm |  | Point | 0.0528 | Unitless |  |
| Dose-response parameter, infection endpoint | r | Lognormal^a^ | μ=-2.93, σ=0.49 | Unitless | Armstrong and Haas ^8^,Muller, et al. ^9^ |
| Attack rate | AR | Point | 0.05 | Unitless | Weir, et al. ^10^ |
| Incidence rate | I |  |  |  | BAG ^11^ |
| Child |  | Point | 1.2e-5 | Unitless |  |
| Adult |  | Point | 4.5e-4 | Unitless |  |
| Elderly |  | Point | 1.4e-3 | Unitless |  |
| Total population |  | Point | 1.9e-3 | Unitless |  |
| Disease severity | DS |  |  | Unitless | Blanky, et al. ^12^ |
| Legionnaire’s disease |  | Point | 0.3 | Unitless |  |
| Disease duration | T |  |  |  | Blanky, et al. ^12^ |
| Legionnaire’s disease |  | Point | 21 | days |  |
| Years of life lost for legionellosis patients | L | Point | 21.8 | years | Blanky, et al. ^12^ |
| Mortality rate of legionnaire’s disease | Mt | Point | 0.08 | Unitless | Zanella, et al. ^13^ |

^a^Lognormal parameters mean, standard deviation (μ, σ) calculated from population (normal) parameters (x̅, s) using standard formulas as follows:μ = ln(x̅2 /(s2 + x̅ 2) 1/2), σ = [ln(1 + (s2 /x̅2 ))] 1/2, where x̅ is the sample mean and s is the sample standard deviation.

**Table S2**. Concentrations of *L. pneumophila* sampled from the studied at-risk building

| Shower ID | Sampling date | Sample type | Water temperature | Concentration (CFU/L) |
| --- | --- | --- | --- | --- |
| Shower A | 03.03.2022 | First draw | Cold | 20 |
| Shower A | 03.03.2022 | Flushed | Cold | 180 |
| Shower A | 03.03.2022 | First draw | Hot | 40 |
| Shower A | 03.03.2022 | Flushed | Hot | 20 |
| Shower A | 08.02.2022 | First draw | Cold | 8000 |
| Shower A | 08.02.2022 | Flushed | Cold | 2260 |
| Shower A | 08.02.2022 | First draw | Hot | 20000 |
| Shower A | 08.02.2022 | Flushed | Hot | 4080 |
| Shower A | 12.10.2022 | First draw | Cold | 1600 |
| Shower A | 12.10.2022 | Flushed | Cold | 440 |
| Shower A | 12.10.2022 | First draw | Hot | 40 |
| Shower A | 12.10.2022 | Flushed | Hot | 100 |
| Shower A | 14.09.2022 | First draw | Cold | 1000 |
| Shower A | 14.09.2022 | Flushed | Cold | 480 |
| Shower A | 14.09.2022 | First draw | Hot | 60 |
| Shower A | 14.09.2022 | Flushed | Hot | 0 |
| Shower A | 17.08.2022 | First draw | Cold | 280 |
| Shower A | 17.08.2022 | Flushed | Cold | 200 |
| Shower A | 17.08.2022 | First draw | Hot | 1500 |
| Shower A | 17.08.2022 | Flushed | Hot | 500 |
| Shower A | 19.04.2022 | First draw | Cold | 1720 |
| Shower A | 19.04.2022 | Flushed | Cold | 800 |
| Shower A | 19.04.2022 | First draw | Hot | 60 |
| Shower A | 19.04.2022 | Flushed | Hot | 60 |
| Shower A | 20.05.2022 | First draw | Cold | 1920 |
| Shower A | 20.05.2022 | Flushed | Cold | 440 |
| Shower A | 20.05.2022 | First draw | Hot | 300 |
| Shower A | 20.05.2022 | Flushed | Hot | 400 |
| Shower A | 25.03.2022 | First draw | Cold | 940 |
| Shower A | 25.03.2022 | Flushed | Cold | 380 |
| Shower A | 25.03.2022 | First draw | Hot | 40 |
| Shower A | 25.03.2022 | Flushed | Hot | 40 |
| Shower A | 28.10.2021 | First draw | Cold | 19000 |
| Shower A | 28.10.2021 | Flushed | Cold | 7000 |
| Shower A | 28.10.2021 | First draw | Hot | 7000 |
| Shower A | 28.10.2021 | Flushed | Hot | 5000 |
| Shower B | 28.10.2021 | First draw | Cold | 59000 |
| Shower B | 28.10.2021 | Flushed | Cold | 19000 |
| Shower B | 28.10.2021 | First draw | Hot | 25000 |
| Shower B | 28.10.2021 | Flushed | Hot | 14000 |
| Shower C | 28.10.2021 | First draw | Cold | 13000 |
| Shower C | 28.10.2021 | Flushed | Cold | 11000 |
| Shower C | 28.10.2021 | First draw | Hot | 7000 |
| Shower C | 28.10.2021 | Flushed | Hot | 5000 |
| Shower D | 03.03.2022 | First draw | Cold | 300 |
| Shower D | 03.03.2022 | Flushed | Cold | 40 |
| Shower D | 03.03.2022 | First draw | Hot | 80 |
| Shower D | 03.03.2022 | Flushed | Hot | 40 |
| Shower D | 08.02.2022 | First draw | Cold | 43000 |
| Shower D | 08.02.2022 | Flushed | Cold | 9000 |
| Shower D | 08.02.2022 | First draw | Hot | 3000 |
| Shower D | 08.02.2022 | Flushed | Hot | 10200 |
| Shower D | 12.10.2022 | First draw | Cold | 51000 |
| Shower D | 12.10.2022 | Flushed | Cold | 580 |
| Shower D | 12.10.2022 | First draw | Hot | 1000 |
| Shower D | 12.10.2022 | Flushed | Hot | 740 |
| Shower D | 14.09.2022 | First draw | Cold | 7000 |
| Shower D | 14.09.2022 | Flushed | Cold | 220 |
| Shower D | 14.09.2022 | First draw | Hot | 1000 |
| Shower D | 14.09.2022 | Flushed | Hot | 280 |
| Shower D | 17.08.2022 | First draw | Cold | 26000 |
| Shower D | 17.08.2022 | Flushed | Cold | 400 |
| Shower D | 17.08.2022 | First draw | Hot | 2000 |
| Shower D | 17.08.2022 | Flushed | Hot | 440 |
| Shower D | 19.04.2022 | First draw | Cold | 29000 |
| Shower D | 19.04.2022 | Flushed | Cold | 5000 |
| Shower D | 19.04.2022 | First draw | Hot | 1740 |
| Shower D | 19.04.2022 | Flushed | Hot | 580 |
| Shower D | 20.05.2022 | First draw | Cold | 8000 |
| Shower D | 20.05.2022 | Flushed | Cold | 1140 |
| Shower D | 20.05.2022 | First draw | Hot | 680 |
| Shower D | 20.05.2022 | Flushed | Hot | 1060 |
| Shower D | 25.03.2022 | First draw | Cold | 1260 |
| Shower D | 25.03.2022 | Flushed | Cold | 540 |
| Shower D | 25.03.2022 | First draw | Hot | 420 |
| Shower D | 25.03.2022 | Flushed | Hot | 200 |
| Shower D | 28.10.2021 | First draw | Cold | 20000 |
| Shower D | 28.10.2021 | Flushed | Cold | 16000 |
| Shower D | 28.10.2021 | First draw | Hot | 10000 |
| Shower D | 28.10.2021 | Flushed | Hot | 7000 |
| Shower E | 03.03.2022 | First draw | Cold | 560 |
| Shower E | 03.03.2022 | Flushed | Cold | 100 |
| Shower E | 03.03.2022 | First draw | Hot | 60 |
| Shower E | 03.03.2022 | Flushed | Hot | 40 |
| Shower E | 08.02.2022 | First draw | Cold | 7000 |
| Shower E | 08.02.2022 | Flushed | Cold | 1360 |
| Shower E | 08.02.2022 | First draw | Hot | 0 |
| Shower E | 08.02.2022 | Flushed | Hot | 180 |
| Shower E | 12.10.2022 | First draw | Cold | 1400 |
| Shower E | 12.10.2022 | Flushed | Cold | 1200 |
| Shower E | 12.10.2022 | First draw | Hot | 180 |
| Shower E | 12.10.2022 | Flushed | Hot | 140 |
| Shower E | 14.09.2022 | First draw | Cold | 5000 |
| Shower E | 14.09.2022 | Flushed | Cold | 800 |
| Shower E | 14.09.2022 | First draw | Hot | 300 |
| Shower E | 14.09.2022 | Flushed | Hot | 280 |
| Shower E | 17.08.2022 | First draw | Cold | 6000 |
| Shower E | 17.08.2022 | Flushed | Cold | 6000 |
| Shower E | 17.08.2022 | First draw | Hot | 940 |
| Shower E | 17.08.2022 | Flushed | Hot | 300 |
| Shower E | 19.04.2022 | First draw | Cold | 5000 |
| Shower E | 19.04.2022 | Flushed | Cold | 700 |
| Shower E | 19.04.2022 | First draw | Hot | 700 |
| Shower E | 19.04.2022 | Flushed | Hot | 20 |
| Shower E | 20.05.2022 | First draw | Cold | 1040 |
| Shower E | 20.05.2022 | Flushed | Cold | 440 |
| Shower E | 20.05.2022 | First draw | Hot | 180 |
| Shower E | 20.05.2022 | Flushed | Hot | 0 |
| Shower E | 25.03.2022 | First draw | Cold | 600 |
| Shower E | 25.03.2022 | Flushed | Cold | 240 |
| Shower E | 25.03.2022 | First draw | Hot | 200 |
| Shower E | 25.03.2022 | Flushed | Hot | 20 |
| Shower E | 28.10.2021 | First draw | Cold | 37000 |
| Shower E | 28.10.2021 | Flushed | Cold | 8000 |
| Shower F | 03.03.2022 | First draw | Cold | 7000 |
| Shower F | 03.03.2022 | Flushed | Cold | 140 |
| Shower F | 03.03.2022 | First draw | Hot | 60 |
| Shower F | 03.03.2022 | Flushed | Hot | 0 |
| Shower F | 08.02.2022 | First draw | Cold | 9000 |
| Shower F | 08.02.2022 | Flushed | Cold | 1380 |
| Shower F | 08.02.2022 | First draw | Hot | 4080 |
| Shower F | 08.02.2022 | Flushed | Hot | 140 |
| Shower F | 12.10.2022 | First draw | Cold | 5000 |
| Shower F | 12.10.2022 | Flushed | Cold | 1800 |
| Shower F | 12.10.2022 | First draw | Hot | 40 |
| Shower F | 12.10.2022 | Flushed | Hot | 40 |
| Shower F | 14.09.2022 | First draw | Cold | 5000 |
| Shower F | 14.09.2022 | Flushed | Cold | 500 |
| Shower F | 14.09.2022 | First draw | Hot | 500 |
| Shower F | 14.09.2022 | Flushed | Hot | 0 |
| Shower F | 17.08.2022 | First draw | Cold | 7000 |
| Shower F | 17.08.2022 | Flushed | Cold | 5000 |
| Shower F | 17.08.2022 | First draw | Hot | 180 |
| Shower F | 17.08.2022 | Flushed | Hot | 0 |
| Shower F | 19.04.2022 | First draw | Cold | 21000 |
| Shower F | 19.04.2022 | Flushed | Cold | 16000 |
| Shower F | 19.04.2022 | First draw | Hot | 5000 |
| Shower F | 19.04.2022 | Flushed | Hot | 560 |
| Shower F | 20.05.2022 | First draw | Cold | 6000 |
| Shower F | 20.05.2022 | Flushed | Cold | 1700 |
| Shower F | 20.05.2022 | First draw | Hot | 680 |
| Shower F | 20.05.2022 | Flushed | Hot | 1120 |
| Shower F | 25.03.2022 | First draw | Cold | 4000 |
| Shower F | 25.03.2022 | Flushed | Cold | 1260 |
| Shower F | 25.03.2022 | First draw | Hot | 19000 |
| Shower F | 25.03.2022 | Flushed | Hot | 0 |
| Shower F | 28.10.2021 | First draw | Cold | 28000 |
| Shower F | 28.10.2021 | Flushed | Cold | 28000 |
| Shower F | 28.10.2021 | First draw | Hot | 5000 |
| Shower F | 28.10.2021 | Flushed | Hot | 4000 |
